# Evaluating the Real-World Safety of Atypical Antipsychotics in Paediatric Autism Spectrum Disorder: Lessons from Saudi Arabia’s Pharmacovigilance System

**DOI:** 10.64898/2026.09.22.26363643

**Authors:** Mashal Aljead, Mohammed Almeziny, Eman Alghamdi, Zahraa Jalal, Alan M. Jones

## Abstract

**Background:** Atypical antipsychotics (AAPs) are increasingly prescribed for paediatric patients with autism spectrum disorder (ASD). However, pharmacovigilance data from Saudi Arabia remain scarce. The aim of this study is to detect and characterise signals associated with AAPs and assess gender- and age-related ADR reporting patterns.

**Methods:** A retrospective cross–sectional study with disproportionality analysis was conducted on paediatric patients (0–19 years) using the Saudi Food and Drug Authority (SFDA) database (2021–2024). Disproportionality analyses were performed using the reporting odds ratio (ROR), proportional reporting ratio (PRR), and Bayesian confidence propagation neural network (BCPNN). Age and gender were assessed using odds ratios (ORs), with Firth penalized logistic regression to confirm associations.

**Results:** Overall, 701 ADR reports were analysed. Most involved males (59.91%) and children aged 0-9 years (82.88%). Olanzapine had the highest proportion of serious reports (44.44%). At the system organ class (SOC) level, signals included nervous system disorders with risperidone (ROR 32.61 [16.62–64.00]; PRR 3.49 [2.22–5.47]; IC025 0.08); cardiac and general disorders with quetiapine; haematological, metabolic, and psychiatric disorders with olanzapine; and gastrointestinal disorders with aripiprazole. Gender–adjusted analyses indicated significant variation in selected ADR signals across age groups, while no significant differences were observed by gender.

**Conclusions:** This study highlights disproportionality signals associated with AAP use in paediatric patients. Age–related ADR variation supports consideration of age-specific ADR monitoring within current paediatric recommendations. While these findings do not establish causality, they generate hypotheses for further evaluation. Regional pharmacovigilance may complement international paediatric safety monitoring frameworks.

## 1. Introduction

Over the last 10 years, the prescribing of atypical antipsychotics (AAPs) for paediatric patients with autism spectrum disorder (ASD) has increased globally, with a pooled prevalence of 5.5 per 1,000 across 23 countries (Downey et al., 2026; Piovani et al., 2019). This increase may reflect their established efficacy in controlling associated behavioural symptoms (Libowitz and Nurmi, 2021; Panda et al., 2025). Risperidone and aripiprazole are prescribed as first-line of AAPs in several international guidelines. These include the practice parameter for the use of atypical antipsychotic medications in children and adolescents in the United States (US), the Maudsley prescribing guidelines in the United Kingdom (UK), and the Saudi Arabian guidelines for ASD treatment and monitoring (Aljead et al., 2025b; D’Alò et al., 2021; Dinnissen et al., 2020). Other AAPs, namely olanzapine, quetiapine, lurasidone, and ziprasidone, are administered off-label in cases of failure of the first-line treatment of AAPs (Lopes et al., 2026). Despite their clinical benefits, serious adverse drug reactions (ADRs) continue to be reported through spontaneous reporting systems (SRS) and observational studies. SRSs such as the Food and Drug Administration (FDA) Adverse Event Reporting System (FAERS), the Medicines and Healthcare products Regulatory Agency (MHRA) Yellow Card scheme, and the Saudi Food and Drug Authority (SFDA) have increased awareness about ADRs associated with medications *via* post-marketing surveillance (Aljead et al., 2025a). Although these systems have several limitations, including a lack of reliable prevalence data on ADRs and critical data such as clinical indication, dose, and drug-drug interactions, as well as underreporting, they provide important post-marketing safety data to support drug safety monitoring (Dedefo et al., 2025). Moreover, SRSs can detect rare or long latency ADRs that clinical trials do not capture (Gavin et al., 2025; Loke et al., 2024).

Despite the growing use of AAPs in paediatric patients with ASD, pharmacovigilance studies from the Middle East remain limited (Caster et al., 2020). Most published disproportionality analyses have been conducted using Western databases, particularly the MHRA and FAERS, with limited representation of regional populations who’s genetic and healthcare system differences may influence ADR reporting (Cepaityte et al., 2021; Gui et al., 2025). The SFDA database offers a valuable yet underutilized resource for examining ADR patterns in Saudi Arabia, which had an estimated population of 34 million in 2023, where ASD prevalence is estimated at 2.5% (Alenzi et al., 2022; Alsulami, 2025; Yee et al., 2026).

The aim of this study is to detect and characterise signals of disproportionate reporting associated with AAP use in paediatric patients with ASD using the SFDA database and to assess gender and age as factors associated with ADR reporting patterns at both the system organ class (SOC) and preferred term (PT) levels.

## 2. Methods

### Data source and processing

A retrospective, cross-sectional, descriptive analysis with disproportionality analysis, which was conducted according to READUS-PV (REporting of A Disproportionality analysis for drUg Safety signal detection using individual case safety reports in PharmacoVigilance) guidelines (Fusaroli et al., 2024). Disproportionality analysis was selected because it is a recognised approach for detecting potential safety signals in SRSs when prevalence and incidence rates cannot be estimated (Fusaroli et al., 2026; Zhou et al., 2025). This study analysed data extracted from the SFDA database from January 2021 to December 2024, for paediatric patients (0–19 years) who were prescribed AAPs. According to the World Health Organization (WHO) definition of the paediatric population, age was categorised into two groups: children (0-9 years) and adolescents (10-19 years) (Organization, 2024). A total of 721 reports were initially identified in the SFDA. After removing duplicate reports (2 reports), incomplete data (6 reports), and reports not involving the target medications (12 reports), 701 ADR reports were included in the final analysis. Reports submitted by health care professionals (HCPs), patients, and medication manufacturers were included. Each report is termed an Individual Case Safety Report (ICSR) and is coded using Medical Dictionary for Regulatory Activities (MedDRA) at the System Organ Class (SOC) and preferred term (PT) levels. Each report included the following: ADR (PT), age, gender, drug name, ADR (SOC), and seriousness.

### Ethical Approval

The study used anonymized ICSRs obtained from the SFDA pharmacovigilance database following ethical approval from the SFDA (No. 2025_016) and the Science, Technology, Engineering, and Mathematics Ethics Committee at the University of Birmingham (ERN_4166-May2025), with regulatory permission from the SFDA.

### Data management and statistical analysis

A two-by-two contingency table was created as shown in Table 1, which forms the basis for all statistics in the disproportionate analysis and factors associated with ADR reporting patterns.

**Table 1.** Two-by-two contingency table. **a:** number of reports for drug X with adverse drug reaction Y; **b:** number of reports for drug X with other adverse drug reactions; **c:** number of reports for all other drugs with adverse drug reaction Y; **d:** number of reports for all other drugs with other adverse drug reactions.

|  | Adverse event Y | All other adverse events | Total |
| --- | --- | --- | --- |
| Reports for drug of interest (X) | a | b | a + b |
| All Other Drugs | c | d | c + d |
| Total | a + c | b + d | a + b + c + d |

Subsequently, signal detection was performed by calculating the reporting odds ratio (ROR), the proportional reporting ratio (PRR), and a Bayesian confidence propagation neural network (BCPNN) to calculate the information component (IC). Positive signal detection was confirmed if three measurements met the following criteria:

- ROR ([(a/c)/(b/d)]): ROR ≥ 2, a ≥ 3, and 95% CI (lower limit) > 1
- PRR [ a/(a+b)] / [c/(c+d)]: PRR ≥ 2, a ≥ 3, and 95% CI (lower limit) > 1
- IC = log2[(a + 0.5)/ (()a + b) (a + c))/(a + b + c + d)) + 0.5)]: IC025 > 0 and a ≥ 3

When a zero-cell occurred in the contingency table, the Haldane-Anscombe correction was applied(Sun et al., 2025).

For categorical comparisons, the chi-square was used, with Fisher’s exact test applied when any expected cell count was <5. Gender and age were assessed as factors associated with ADR reporting patterns using odds ratios (ORs), with Firth penalized logistic regression applied to confirm associations. For these analyses, A *p*-value < 0.05 and a 95% confidence interval (CI) that does not cross 1.0 were considered statistically significant.

Microsoft Excel (version 2605)was used for curating, cleaning, and filtering data according to predefined inclusion criteria. All statistical analyses were conducted using *R* software (version 4.5.2).

## 3. Results

Descriptive demographic and clinical characteristics of 701 ADR reports identified in the SFDA database between January 2021and December 2024 are presented in Table 2. Risperidone accounted for the majority of reports (92.44%), followed by olanzapine (3.85%), quetiapine (3.00%), and aripiprazole (0.71%). Most reports involved males (59.91%) and children aged 0-9 years (82.88%). Although the majority of ADRs were classified as non-serious (95.57%), olanzapine accounted for the highest proportion of serious ADR reports (44.44%).

**Table 2.** Demographic distribution by gender, age group, and seriousness of SFDA spontaneous reports in paediatric patients with autism spectrum disorder (ASD) associated with atypical antipsychotics (AAPs) (2021–2024). Percentages were calculated using the relevant column as the denominator (Total: n = 701; risperidone: n = 648; aripiprazole: n = 5; olanzapine: n = 27; quetiapine: n = 21).

|  | **Total** | **Risperidone**<br>**no. (%)** | **Aripiprazole**<br>**no. (%)** | **Olanzapine no.**<br>**(%)** | **Quetiapine**<br>**no. (%)** |
| --- | --- | --- | --- | --- | --- |
| **Number of reports** | 701 | 648 (92.44%) | 5 (0.71%) | 27 (3.85%) | 21 (3.00%) |
| **Male (%)** | 420 (59.91%) | 394 (60.80%) | 2 (40.00%) | 15 (55.56%) | 9 (42.86%) |
| **Female (%)** | 281 (40.09%) | 254 (39.20%) | 3 (60.00%) | 12 (44.44%) | 12 (57.14%) |
| **Children: Age**<br>**group-0-9 Years (%)** | 581 (82.88%) | 572 (88.27%) | 2 (40.00%) | 0 (0%) | 7 (33.33%) |
| **Adolescents: Age**<br>**group-10-19 Years**<br>**(%)** | 120 (17.12%) | 76 (11.73%) | 3 (60.00%) | 27 (100%) | 14 (66.67%) |
| **Non-serious ADR**<br>**(%)** | 670 (95.57%) | 636 (98.15%) | 5 (100%) | 15 (55.56%) | 14 (66.67%) |
| **Serious ADR (%)** | 31 (4.42%) | 12 (1.85%) | 0 (0%) | 12 (44.44%) | 7 (33.33%) |

Results are presented in three levels (1) disproportionality analysis at the SOC (Table 3) and PT levels (Table 4 and Supplementary Table 1) (2) assessment of gender as a factor associated with ADR reporting patterns at the SOC (Supplementary Table 2 and 3) and PT levels (Supplementary Table 4 and 5); and (3) assessment of age as a factor associated with ADR reporting patterns at the SOC (Table 5 and Supplementary Table 3 and 6) and PT levels (Supplementary Table 5 and 7).

**Table 3.** System organ class (SOC)–level disproportionality analysis of adverse drug reaction (ADR) reporting associated with atypical antipsychotics (AAPs) in paediatric autism spectrum disorder (ASD). 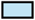Positive Disproportionality Signal: ROR ≥ 2, PRR ≥ 2, a ≥ 3 and 95% CI (lower limit) > 1, IC025 > 0, (–): not calculated (a = 0)

| AAP | Risperidone |  |  |  | Aripiprazole |  |  |  | Olanzapine |  |  |  | Quetiapine |  |  |  |
| --- | --- | --- | --- | --- | --- | --- | --- | --- | --- | --- | --- | --- | --- | --- | --- | --- |
| SOC | a | ROR<br>(95%CI) | PRR<br>(95%CI) | IC<br>(IC025) | a | ROR<br>(95%CI) | PRR<br>(95%CI) | IC<br>(IC025) | a | ROR<br>(95%CI) | PRR<br>(95%CI) | IC<br>(IC025) | a | ROR<br>(95%CI) | PRR<br>(95%CI) | IC<br>(IC025) |
| Blood and lymphatic system disorders | 0 | — | — | — | 0 | — | — | — | 4 | 258.32<br>(13.51–4938.41) | 216.96<br>(11.97–3932.27) | 4.50 (2.59) | 0 | — | — | — |
| Cardiac disorders | 0 | — | — | — | 0 | — | — | — | 0 | — | — | — | 4 | 349.97<br>(18.13–6754.26) | 278.59<br>(15.47–5016.70) | 4.85<br>(2.92) |
| Endocrine disorders | 3 | 0.24<br>(0.02–2.37) | 0.25<br>(0.03–2.32) | -0.30 (-2.46) | 0 | — | — | — | 1 | 8.60<br>(0.87–85.54) | 8.32<br>(0.89–77.41) | 2.70 (-0.50) | 0 | — | — | — |
| Gastrointestinal disorders | 13 | 0.20<br>(0.07–0.57) | 0.21<br>(0.08–0.57) | -0.36 (-1.37) | 3 | 68.10<br>(10.59–437.87) | 27.84<br>(11.62–66.68) | 4.55<br>(2.38) | 0 | — | — | — | 2 | 4.37<br>(0.94–20.36) | 4.05<br>(0.99–16.49) | 1.89 (-0.30) |
| General disorders and administration site conditions | 6 | 0.07<br>(0.02–0.24) | 0.08<br>(0.03–0.24) | -0.89 (-2.29) | 0 | — | — | — | 3 | 9.24<br>(2.35–36.30) | 8.32<br>(2.39–29.01) | 2.70 (0.80) | 3 | 12.43<br>(3.10–49.79) | 10.79<br>(3.15–37.02) | 3.06<br>(1.14) |
| Hepatobiliary disorders | 2 | 0.08<br>(0.01–0.57) | 0.08<br>(0.01–0.57) | -0.89 (-3.33) | 0 | — | — | — | 2 | 26.88<br>(3.64–198.67) | 24.96<br>(3.65–170.59) | 3.70 (1.20) | 0 | — | — | — |
| Metabolism and nutrition disorders | 14 | 0.17<br>(0.06–0.47) | 0.19<br>(0.08–0.48) | -0.40 (-1.38) | 0 | — | — | — | 5 | 9.98<br>(3.33–29.93) | 8.32<br>(3.26–21.22) | 2.70 (1.19) | 1 | 1.74<br>(0.22–13.64) | 1.70<br>(0.24–12.14) | 0.74 (-2.22) |
| Musculoskeletal and connective tissue disorders | 4 | 0.32<br>(0.04–2.94) | 0.33<br>(0.04–2.88) | -0.21 (-2.10) | 0 | — | — | — | 1 | 6.44<br>(0.70–59.68) | 6.24<br>(0.72–53.97) | 2.38 (-0.76) | 0 | — | — | — |
| Nervous system disorders | 597 | 32.61<br>(16.62–64.00) | 3.49<br>(2.22–5.47) | 0.08<br>(0.01) | 2 | 0.10<br>(0.02–0.58) | 0.46<br>(0.16–1.34) | -1.12 (-3.49) | 3 | 0.01<br>(0.00–0.05) | 0.12<br>(0.04–0.36) | -2.97 (-4.69) | 9 | 0.10<br>(0.04–0.24) | 0.48<br>(0.30–0.79) | -1.02 (-2.14) |
| Psychiatric disorders | 5 | 0.07<br>(0.02–0.27) | 0.08<br>(0.02–0.27) | -0.89 (-2.43) | 0 | — | — | — | 5 | 30.41<br>(8.20–112.74) | 24.96<br>(7.68–81.11) | 3.70 (2.07) | 0 | — | — | — |
| **Renal and urinary disorders** | 1 | 0.25<br>(0.01–6.16) | 0.25<br>(0.01–6.05) | -0.30 (-3.35) | 0 | – | – | – | 0 | – | – | – | 0 | – | – | – |
| **Reproductive system and breast disorders** | 0 | – | – | – | 0 | – | – | – | 0 | – | – | – | 1 | 99.59<br>(3.94–2519.20) | 92.86<br>(3.89–2216.12) | 4.58<br>(1.48) |
| **Respiratory, thoracic and mediastinal disorders** | 0 | – | – | – | 0 | – | – | – | 1 | 76.36<br>(3.04–1918.99) | 72.32<br>(3.01–1736.08) | 4.23 (1.14) | 0 | – | – | – |
| **Skin and subcutaneous tissue disorders** | 3 | 0.12<br>(0.02–0.73) | 0.12<br>(0.02–0.72) | -0.62 (-2.68) | 0 | – | – | – | 2 | 17.89<br>(2.86–111.91) | 16.64<br>(2.90–95.52) | 3.38 (0.96) | 0 | – | – | – |
| **Vascular disorders** | 0 | – | – | – | 0 | – | – | – | 0 | – | – | – | 1 | 99.59<br>(3.94–2519.20) | 92.86<br>(3.89–2216.12) | 4.58<br>(1.48) |

**Table 4.** Detection of positive disproportionality signals for selected adverse drug reactions (ADRs) associated with atypical antipsychotics (AAPs).

| AAP | PT | a | ROR (95%CI) | PRR (95%CI) | IC (IC025) |
| --- | --- | --- | --- | --- | --- |
| Olanzapine | Neutropenia | 4.00 | 258.32 (13.51–4938.41) | 216.96 (11.97–3932.27) | 4.49 (2.59) |
| Quetiapine | Electrocardiogram QT prolonged | 4.00 | 349.97 (18.13–6754.26) | 278.59 (15.47–5016.70) | 4.84(2.92) |

**Table 5.** Age-related differences in SOC-level ADR reporting patterns in paediatric patients with ASD associated with atypical antipsychotics (AAPs): Firth penalized logistic regression analysis (aOR, Adjusted OR, 95% CI).

| AAP | SOC | Age 10–19 ADR no. | Age 0–9 ADR no. | Univariate analysis |  | Multivariate analysis |  |
| --- | --- | --- | --- | --- | --- | --- | --- |
|  |  |  |  | OR (95% CI) | P value | aOR (95% CI) | Adjusted P value |
| Risperidone | Gastrointestinal disorders | 5 | 8 | 4.96 (1.58–15.59) | 0.01 | 5.46 (1.69–16.31) | 0.01 |
| Risperidone | General disorders and administration site conditions | 4 | 2 | 15.83 (2.85–87.98) | <0.001 | 14.97 (3.23–87.84) | <0.001 |
| Risperidone | Musculoskeletal and connective tissue disorders | 3 | 1 | 23.47 (2.41–228.57) | 0.01 | 17.42 (2.81–181.95) | <0.001 |
| Risperidone | Nervous system disorders | 56 | 541 | 0.16 (0.09–0.30) | <0.001 | 0.15 (0.08–0.29) | <0.001 |
| Risperidone | Renal and urinary disorders | 1 | 0 | 22.75 (0.92–563.49) | 0.01 | 20.79 (1.09–3055.9) | 0.04 |
| Risperidone | Skin and subcutaneous tissue disorders | 2 | 1 | 15.43 (1.38–172.29) | 0.04 | 12.64 (1.65–140.18) | 0.02 |

### Disproportionality analysis

Risperidone accounted for the largest proportion of reports ( n = 648). Compared with risperidone; aripiprazole (n = 5), olanzapine ( n =27), and quetiapine (n =21) had fewer reports with wider confidence intervals. This resulted in imprecise disproportionality signals (Table 3).

Several positive disproportionality signals were observed across AAPs (Table 3). Risperidone demonstrated a positive disproportionality signal for nervous system disorders, with ROR 32.61 (95% CI 16.62–64.00), PRR 3.49 (95% CI 2.22– 5.47), and IC 0.08 (IC025 0.01). Aripiprazole showed a positive signal for gastrointestinal disorders, with ROR 68.10 (10.59–437.87), PRR 27.84 (11.62– 66.68), and IC 4.55 (IC025 2.38).

By contrast, olanzapine showed multiple positive signals, including blood and lymphatic disorders, general disorders and administration site conditions, metabolism and nutrition disorders, and psychiatric disorders. The strongest signal was observed for metabolism and nutrition disorders, with ROR 9.98 (95% CI 3.33–29.93), PRR 8.32 (95% CI 3.62–21.22), and IC 2.70 (IC025 1.19) (Table 3). For quetiapine, two positive disproportionality signals were detected with cardiac disorders and general disorders and administration site conditions. The strongest signal was observed for cardiac disorders, ROR 349.97 (95% CI 18.13– 6754.26), PRR 278.59 (95% CI 15.47–5016.70), and IC 4.85 (IC025 2.92) (Table 3).

The signals for blood and lymphatic disorders with olanzapine and cardiac disorders with quetiapine were driven exclusively by neutropenia and electrocardiogram QT prolonged, respectively (Table 4). The complete PT-level disproportionality analysis is presented in Supplementary Table 1.

### Gender differences

Gender-stratified analyses compared females with males (male reference). At the SOC level, no significant gender differences in ADR reporting patterns were observed across SOC categories for any drug (Supplementary Table 2).

After adjusting for age, the Firth penalized logistic regression analysis did not identify any significant associations between gender and SOC categories (Supplementary Table 3).

Similarly, no statistically significant gender-related differences in ADR reporting patterns were identified in PT-level analyses in either gender-stratified analyses (Supplementary Table 4) or age-adjusted analyses (Supplementary Table 5).

### Age differences

Age-stratified analyses compared adolescents (10-19 years) with children aged 0-9 years (reference group). At the SOC level, after adjusting for gender using Firth penalized logistic regression, the associations remained or became significant only with risperidone (Table 5). Adolescents had significantly higher adjusted odds of gastrointestinal disorders (aOR 5.46, 95% CI 1.69–16.31, P = 0.01), general disorders and administration site conditions (aOR 14.97, 95% CI 3.23– 87.84, P < 0.001), musculoskeletal and connective tissue disorders (aOR 17.42, 95% CI 2.81–181.95, P < 0.001), renal and urinary disorders (aOR 20.79, 95% CI 1.09–3055.9, P = 0.04), and skin and subcutaneous tissue disorders (aOR 12.64, 95% CI 1.65–140.18, P = 0.02) compared with children.

Conversely, adolescents had significantly lower adjusted odds of nervous system disorders (aOR 0.15, 95% CI 0.08–0.29, P < 0.001). Full SOC-level, age-stratified and gender-adjusted analyses across AAPs are presented in Supplementary Tables 3 and 6, respectively.

Similarly, no statistically significant age-related differences in ADR reporting patterns were identified at the PT-level in either the age-stratified (Supplementary Table 7) or the gender-adjusted analyses (Supplementary Table 5).

## 4. Discussion

This study is the first pharmacovigilance analysis to provide an overview of disproportionality signals and to assess gender and age-related differences in ADR reporting patterns associated with AAPs in children and adolescents with ASD from the SFDA secure data environment. Using multiple disproportionality measures and stringent criteria, the study prioritised signals that met predefined signal-detection criteria and may warrant further clinical evaluation and monitoring.

A key methodological strength is integration of SOC-level and PT-level analyses. SOC analyses increase sensitivity by aggregating related PTs, which is useful when PT counts are small. Meanwhile, PT analyses increase specificity and can clarify the nature of a SOC signal. For example, blood and lymphatic disorders and cardiac disorders signals were driven exclusively by neutropenia and QT prolongation, respectively.

The findings indicate that most reports were reported in males and associated with risperidone. This pattern may reflect prescribing practices, particularly the more frequent use of risperidone as a first-line treatment for irritability associated ASD (Alsabhan et al., 2024; Egberts et al., 2022). Additionally, the higher prevalence of ASD diagnoses in males may partly explain the higher proportion of reports among males, rather than comparative drug safety profiles alone (Aljead et al., 2025b).

Several positive disproportionality signals were identified at the SOC level across different AAPs. Olanzapine demonstrated positive disproportionality signals in multiple SOCs, including blood and lymphatic system disorders, metabolism and nutrition disorders, and psychiatric disorders. The blood and lymphatic disorders signal was primarily driven by neutropenia, which is a recognised ADR that may be related to bone marrow suppression (Min and Byeon, 2025). Although SRS cannot establish causality, the concordance between SOC-level statistical signals and PT-level qualitative findings may increase confidence in the clinical relevance of this signal. These findings may support further evaluation of haematological safety associated with olanzapine because this serious ADR that may lead to treatment interruption and require enhanced laboratory monitoring (Rast et al., 2025).

In contrast, the metabolic and nutrition disorders signal is pharmacologically plausible because these ADRs may reflect the pharmacological activity of olanzapine through histamine (H_1_) and muscarinic (M_1_) receptor antagonism, leading to increased appetite and metabolic disturbance (Levi et al., 2026; Oruch et al., 2025).

Psychiatric disorders, particularly panic attacks and mania, were identified as unlabelled ADRs because they were not reported in the information of FDA, EMA, and SFDA-labelled products. However, these ADRs have been described in several case reports (Fitz-Gerald et al., 1999; Henry and Demotes-Mainard, 2002; Mandalos and Szarek, 1999). Notably, one published case report described olanzapine associated with mania in a 16-year-old male with ASD (London, 1998). This may support the clinical relevance of this finding despite the limited number of reports in the current study. Importantly, psychiatric ADRs may overlap with behavioural symptoms in children or adolescents with ASD, as well as negatively affect the quality of life (QoL) and difficulties in handling care for ASD patients by caregivers. Therefore, careful baseline behavioural assessment before and after initiation of AAPs, particularly with olanzapine, is recommended (Besag et al., 2026; Saudin et al., 2025).

Moreover, quetiapine showed a positive disproportionality signal for cardiac disorders and general disorders and administration site conditions. The concordance between SOC-level and PT-level findings suggests that QT prolongation accounts for the observed disproportionality signal for cardiac ADRs with quetiapine. The QT prolongation signal is clinically significant because it can contribute to serious arrhythmias, although the risk in paediatric populations is considered low (Højlund et al., 2022). Because SRSs lack detailed clinical information, including concomitant QT-prolonging medications (e.g., azithromycin), electrolyte disturbances, and baseline cardiac risk factors, this finding should be considered hypothesis-generating (Hutchins et al., 2021; Jensen et al., 2015). However, the wide confidence interval indicates statistical imprecision and should be interpreted with caution.

Clinically, signals in general disorders and administration site conditions with quetiapine and olanzapine may still be important because they may reflect the overall burden of ADRs and increase caregiver awareness, potentially contributing to increased ADR reporting (Güner et al., 2026).

Aripiprazole demonstrated a positive disproportionality signal for gastrointestinal disorders in this database. This may be due to the reduction of mitochondrial membrane potential and increased oxidative stress in gastrointestinal tissues (Hurcomb et al., 2025). Although biological mechanisms cannot be confirmed using spontaneous reporting data, gastrointestinal ADRs may contribute to treatment interruption and reduced adherence. These ADRs may warrant supportive interventions (e.g., dietary counselling) to prevent treatment interruption and enhance patient adherence (Forman and Zhang, 2021; Hurcomb et al., 2025).

Risperidone accounted for the largest number of ADR reports and demonstrated a strong signal for nervous system disorders. Risperidone is commonly prescribed as first-line therapy for irritability symptoms in children and adolescents with ASD, which may partially explain the larger number of reports (Panda et al., 2025; Yang et al., 2025). Disproportionality analysis compares the proportion of ADR reports rather than the number of reports. Therefore, the nervous system disorders signal is unlikely to be explained by the common use of risperidone alone. Generally, neurological ADRs may substantially affect quality of life and educational performance (Gebru et al., 2025). Somnambulation was identified as an unlabelled ADR associated with risperidone. However, it did not show a positive disproportionality signal, although it has previously been described in several case reports (Detweiler, 2013; Najmi et al., 2020)

In hepatobiliary disorders, musculoskeletal and connective tissue disorders, and skin and subcutaneous tissue disorders, there were a small number of reports of olanzapine and risperidone. Therefore, these findings should not be interpreted as evidence of protective effects or a confirmed increased risk and should be interpreted with caution. Although these ADRs are not serious, events such as alopecia may lead to treatment interruption. Thus, children or adolescents with ASD may require monitoring, as alopecia is more likely in those with ASD than in those without. This may be due to abnormal autoimmune activity and nutritional deficiencies(e.g., zinc) in patients with ASD (Lee et al., 2023).

In renal and urinary disorders, reproductive system and breast disorders, respiratory, thoracic, and mediastinal disorders, and vascular disorders, these findings cannot be considered a robust or confirmatory safety signal because their findings were derived from a single report and should be interpreted with caution.

These findings highlight that different AAPs may exhibit distinct safety profiles across SOCs in paediatric ASD. However, disproportionality analyses reflect the reporting patterns within the database rather than the absolute frequency or risk of ADRs. Consequently, some ADRs may appear over-represented due to reporting background patterns, such as notoriety bias (Cutroneo et al., 2023; Hammad et al., 2025). Therefore, signals were cautiously interpreted and emphasised only when all criteria were met.

Gender differences were generally not robust in this database at either the SOC or PT level. No statistically significant associations were identified after age adjustment, although several non-significant trends were observed. The absence of a statistically significant gender difference might result from a limited number of reports, especially for quetiapine, olanzapine, and aripiprazole. Furthermore, because ASD occurs approximately three times more often in males than in females, this could have influenced the reporting distribution (Aljead et al., 2025b).

Age differences were evident only for risperidone at the SOC level. Adolescents showed higher odds in gastrointestinal, general disorders, musculoskeletal, and skin disorders but lower odds of nervous system disorders compared with children. These associations remained significant after adjustment for gender. In contrast, no significant age-related differences were identified at the PT level. However, the limited number of reports suggests that clinically relevant gender differences cannot be excluded. While age results should be considered exploratory and prioritised for validation using a larger database or clinical cohort, which may guide the age-tailored guideline.

### Strengths and Limitations

This study has several strengths, including being the first to use multiple disproportionality measures, evaluate ADR reporting patterns at both SOC and PT levels, and apply stratified and multivariate modelling to examine age and gender. Furthermore, the focus on paediatric ASD in underrepresented populations such as Saudi Arabia emphasises the importance of addressing this issue in areas with limited resources. However, SRSs are associated with missing data, reporting bias, and under-reporting. Until recently (2020), there has been a lack of guidelines for national databases with 500 or fewer reports, which poses a challenge for robust and clinically relevant signal detection (Caster et al., 2020). Additionally, several ADRs were reported in small numbers and should therefore be considered hypothesis-generating, with priority given to validation in larger databases. Furthermore, the denominator is not available, so the prevalence of ADRs cannot be estimated (Yin et al., 2025). In addition, limited information on medication history, including clinical indication, doses, treatment duration, and concomitant medications, restricts assessment of potential factors (Crisafulli et al., 2025).

### 4.2 Clinical Implications

Although findings from the SRSs cannot establish causality, this study identified several clinically relevant ADR reporting patterns associated with AAPs in paediatric ASD. These findings may support careful baseline and follow-up monitoring tailored to the safety profile of each AAP, particularly in paediatric patients with ASD who may have difficulty recognizing or reporting ADRs. These findings may also support the development of age-specific monitoring recommendations and inform future updates to paediatric ASD treatment and monitoring guidelines in Saudi Arabia.

## 5. Conclusions

This pharmacovigilance study identified ADR reporting patterns and disproportionality signals associated with AAP use in paediatric ASD over four years. Olanzapine was associated with the highest number of positive signals at the SOC level. Risperidone accounted for the largest proportion of ADR reports. Gender differences were not statistically significant, while age-related differences were observed at the SOC level for risperidone. Several unlabelled

ADRs were also identified with both olanzapine and risperidone, highlighting the importance of continued pharmacovigilance in this population. Future analyses in larger pharmacovigilance databases or in clinical cohorts with laboratory data would be valuable to evaluate these ADRs more robustly. A cross-country comparison may additionally help determine whether ADR reporting patterns differ across populations and healthcare systems, potentially supporting region-specific monitoring guidelines.

## Abbreviations

AAPs: Atypical Antipsychotics
ADRs: Adverse Drug Reactions
aOR: Adjusted odds ratio
ASD: Autism Spectrum Disorders
BCPNN: Bayesian confidence propagation neural network
CI: Confidence interval
D_2_: Dopamine D_2_
EMA: European Medicines Agency
FAERS: FDA Adverse Event Reporting System
FDA: Food and Drug Administration
H_1_: Histamine 1
HCPs: Health care professionals
IC: Information component
IC025: lower limit of a 95% credibility interval for the Information component
ICSR: Individual Case Safety Report
M_1_: Muscarinic 1
MedDRA: Medical Dictionary for Regulatory Activities
ME: Middle East
MHRA: The Medicines and Healthcare products Regulatory Agency
NICE: National Institute for Health and Care Excellence
ORs: Odds ratio
PT: Preferred term
PRR: Proportional reporting ratio
QoL: Quality of Life
READUS-PV: REporting of A Disproportionality analysis for drUg Safety signal detection using individual case safety reports in PharmacoVigilance
ROR: Reporting odds ratio
SFDA: Saudi Food and Drug Authority
SOC: System Organ Class
SRSs: Spontaneous reporting systems
US: United States
UK: United Kingdom
WHO: World Health Organization

## CRediT authorship contribution statement

**Mashal Aljead:** Conceptualization, Methodology, Investigation, Data curation, Formal analysis, Resources, Writing—original draft, Writing—review and editing, Visualization, Project administration.

**Mohammed Almeziny:** Writing—review and editing.

**Eman Alghamdi:** Writing—review and editing.

**Zahraa Jalal:** Validation, Writing—review and editing, Supervision.

**Alan M. Jones:** Validation, Writing—review and editing, Supervision.

## Acknowledgements

The authors would like to thank Mr Martin Man for independently confirming the statistical findings. The authors also acknowledge the University of Birmingham for covering the open-access publication fees.

The views expressed in this paper are those of the author(s) and do not necessarily reflect those of the SFDA or its stakeholders. Guaranteeing the accuracy and the validity of the data is the sole responsibility of the research team.

## Data availability

The data analysed in this study were obtained from the Saudi Food and Drug Authority (SFDA) and are not publicly available due to data access restrictions.

### Ethics approval and consent to participate

Ethical approval for this study was obtained from the Saudi Food and Drug Authority (SFDA) Institutional Review Board (Ethics Approval No. 2025_016) and the Science, Technology, Engineering, and Mathematics Ethics Committee at the University of Birmingham (Application No. ERN_4166-May2025). The study used retrospective, de-identified data from the SFDA spontaneous reporting system, with no direct participant involvement or access to identifiable patient information.

## Consent for publication

Not applicable.

## Supplementary material

**Supplementary Table 1:** PT-level disproportionality analysis of adverse drug reaction (ADR) reporting associated with atypical antipsychotics (AAPs) in paediatric patients with autism spectrum disorder (ASD).

**Supplementary Table 2:** Gender-related differences in SOC-level of ADR reporting associated with atypical antipsychotics (AAPs) in paediatric patients with autism spectrum disorder (ASD)(OR, 95% CI).

**Supplementary Table 3:** Firth penalized logistic regression analysis of SOC-level ADR reporting associated with atypical antipsychotics (AAPs) in paediatric patients with autism spectrum disorders (ASD): adjusted odds ratios (aOR, 95%CI).

**Supplementary Table 4:** Gender-related differences in PT-level ADR reporting associated with atypical antipsychotics (AAPs)in paediatric patients with autism spectrum disorder (ASD) (OR, 95% CI).

**Supplementary Table 5:** Firth penalized logistic regression analysis of PT-level ADR reporting associated with atypical antipsychotics (AAPs) in paediatric with autism spectrum disorders (ASD): (aOR, 95%CI).

**Supplementary Table 6:** Age-related differences in SOC level ADR reporting associated with atypical antipsychotics (AAPs) in paediatric patients with autism spectrum disorder (ASD) (OR, 95% CI).

**Supplementary Table 7:** Age-related differences in PT-level ADR reporting associated with atypical antipsychotics (AAPs) in paeadtric patients with autism spectrum disorders (ASD) (OR, 95% CI).

