## Supplementary File for "Evaluating the Real-World Safety of Atypical Antipsychotics in Paediatric Autism Spectrum Disorder: Lessons from Saudi Arabia’s Pharmacovigilance System"

### Supplementary Table 1: PT-level disproportionality analysis of adverse drug reaction (ADR) reporting associated with atypical antipsychotics (AAPs) in paediatric patients with autism spectrum disorder (ASD).

Positive Disproportionality Signal (ROR ≥ 2, PRR ≥ 2, a ≥ 3 and 95% CI (lower limit) > 1, IC025 > 0)

* Unlabeled ADRs (non-positive signals)

| AAP | Risperidone | | | | Aripiprazole | | | | Olanzapine | | | | Quetiapine | | | |
|---|---|---|---|---|---|---|---|---|---|---|---|---|---|---|---|---|
| **PT** | **a** | **ROR (95%CI)** | **PRR (95%CI)** | **IC (IC025)** | **a** | **ROR (95%CI)** | **PRR (95%CI)** | **IC (IC025)** | **a** | **ROR (95%CI)** | **PRR (95%CI)** | **IC (IC025)** | **a** | **ROR (95%CI)** | **PRR (95%CI)** | **IC (IC025)** |
| **Abdominal distension** | 1 | 0.25 (0.01–6.16) | 0.25 (0.01–6.05) | -0.30 (-3.35) | 0 | 42.15 (1.54–1152.96) | 38.72 (1.75–857.96) | 4.87 (0.26) | 0 | 8.16 (0.33–205.00) | 8.04 (0.33–192.90) | 2.65 (-1.85) | 0 | 10.53 (0.42–266.14) | 10.32 (0.43–246.24) | 2.99 (-1.51) |
| **Abdominal pain** | 3 | 0.58 (0.03–11.38) | 0.58 (0.03–11.13) | -0.08 (-2.14) | 0 | 18.01 (0.83–391.83) | 16.60 (0.96–287.53) | 3.87 (-0.52) | 0 | 3.49 (0.18–69.21) | 3.44 (0.18–65.08) | 1.65 (-2.62) | 0 | 4.50 (0.23–89.89) | 4.42 (0.24–83.04) | 1.99 (-2.28) |
| **Abnormal weight gain** | 8 | 0.32 (0.07–1.54) | 0.33 (0.07–1.50) | -0.21 (-1.54) | 0 | 5.94 (0.31–114.54) | 5.53 (0.36–84.05) | 2.41 (-1.83) | 2 | 6.66 (1.34–32.99) | 6.24 (1.39–28.00) | 2.38 (0.13) | 0 | 1.49 (0.08–26.18) | 1.47 (0.09–24.36) | 0.53 (-3.59) |
| **Alopecia** | 0 | 0.03 (0.00–0.67) | 0.03 (0.00–0.67) | -1.89 (-6.35) | 0 | 42.15 (1.54–1152.96) | 38.72 (1.75–857.96) | 4.87 (0.26) | 1 | 76.36 (3.04–1918.99) | 72.32 (3.01–1736.08) | 4.23 (1.14) | 0 | 10.53 (0.42–266.14) | 10.32 (0.43–246.24) | 2.99 (-1.51) |
| **Back pain** | 2 | 0.41 (0.02–8.73) | 0.42 (0.02–8.56) | -0.15 (-2.56) | 0 | 25.25 (1.08–589.47) | 23.23 (1.24–434.30) | 4.28 (-0.18) | 0 | 4.89 (0.23–104.34) | 4.82 (0.24–98.09) | 2.06 (-2.28) | 0 | 6.31 (0.29–135.51) | 6.19 (0.31–125.17) | 2.41 (-1.94) |
| **Confusional state*** | 0 | 0.02 (0.00–0.34) | 0.02 (0.00–0.34) | -2.47 (-6.79) | 0 | 25.25 (1.08–589.47) | 23.23 (1.24–434.30) | 4.28 (-0.18) | 2 | 132.25 (6.19–2826.48) | 120.54 (5.92–2452.21) | 4.38 (1.91) | 0 | 6.31 (0.29–135.51) | 6.19 (0.31–125.17) | 2.41 (-1.94) |
| **Constipation** | 2 | 0.16 (0.01–1.81) | 0.16 (0.02–1.77) | -0.47 (-3.05) | 1 | 86.75 (6.48–1160.88) | 69.60 (7.46–649.55) | 5.55 (2.05) | 0 | 3.49 (0.18–69.21) | 3.44 (0.18–65.08) | 1.65 (-2.62) | 0 | 4.50 (0.23–89.89) | 4.42 (0.24–83.04) | 1.99 (-2.28) |

| | | | | | | | | | | | | | | | | |
|---|---|---|---|---|---|---|---|---|---|---|---|---|---|---|---|---|
| **Cough** | 0 | 0.03 (0.00–0.67) | 0.03 (0.00–0.67) | -1.89 (-6.35) | 0 | 42.15 (1.54–1152.96) | 38.72 (1.75–857.96) | 4.87 (0.26) | 1 | 76.36 (3.04–1918.99) | 72.32 (3.01–1736.08) | 4.23 (1.14) | 0 | 10.53 (0.42–266.14) | 10.32 (0.43–246.24) | 2.99 (-1.51) |
| **Diarrhea** | 1 | 0.25 (0.01–6.16) | 0.25 (0.01–6.05) | -0.30 (-3.35) | 0 | 42.15 (1.54–1152.96) | 38.72 (1.75–857.96) | 4.87 (0.26) | 0 | 8.16 (0.33–205.00) | 8.04 (0.33–192.90) | 2.65 (-1.85) | 0 | 10.53 (0.42–266.14) | 10.32 (0.43–246.24) | 2.99 (-1.51) |
| **Dizziness** | 11 | 0.21 (0.06–0.69) | 0.22 (0.07–0.68) | -0.33 (-1.45) | 1 | 12.18 (1.28–116.05) | 9.94 (1.60–61.87) | 3.22 (0.05) | 1 | 1.81 (0.23–14.32) | 1.78 (0.24–13.07) | 0.79 (-2.17) | 2 | 5.40 (1.14–25.62) | 4.98 (1.20–20.69) | 2.15 (-0.05) |
| **Dry mouth** | 1 | 0.25 (0.01–6.16) | 0.25 (0.01–6.05) | -0.30 (-3.35) | 0 | 42.15 (1.54–1152.96) | 38.72 (1.75–857.96) | 4.87 (0.26) | 0 | 8.16 (0.33–205.00) | 8.04 (0.33–192.90) | 2.65 (-1.85) | 0 | 10.53 (0.42–266.14) | 10.32 (0.43–246.24) | 2.99 (-1.51) |
| **Electrocardiogram QT prolonged** | 0 | 0.01 (0.00–0.16) | 0.01 (0.00–0.17) | -3.21 (-7.40) | 0 | 13.99 (0.67–292.43) | 12.91 (0.78–214.20) | 3.55 (-0.80) | 0 | 2.71 (0.14–51.58) | 2.68 (0.15–48.55) | 1.32 (-2.90) | 4 | 349.97 (18.13–6754.26) | 278.59 (15.47–5016.70) | 4.84 (2.92) |
| **Endocrine disorder** | 1 | 0.25 (0.01–6.16) | 0.25 (0.01–6.05) | -0.30 (-3.35) | 0 | 42.15 (1.54–1152.96) | 38.72 (1.75–857.96) | 4.87 (0.26) | 0 | 8.16 (0.33–205.00) | 8.04 (0.33–192.90) | 2.65 (-1.85) | 0 | 10.53 (0.42–266.14) | 10.32 (0.43–246.24) | 2.99 (-1.51) |
| **Erythema** | 1 | 0.25 (0.01–6.16) | 0.25 (0.01–6.05) | -0.30 (-3.35) | 0 | 42.15 (1.54–1152.96) | 38.72 (1.75–857.96) | 4.87 (0.26) | 0 | 8.16 (0.33–205.00) | 8.04 (0.33–192.90) | 2.65 (-1.85) | 0 | 10.53 (0.42–266.14) | 10.32 (0.43–246.24) | 2.99 (-1.51) |
| **Extrapyramidal disorder** | 3 | 0.24 (0.02–2.37) | 0.25 (0.03–2.32) | -0.30 (-2.46) | 0 | 13.99 (0.67–292.43) | 12.91 (0.78–214.20) | 3.55 (-0.80) | 0 | 2.71 (0.14–51.58) | 2.68 (0.15–48.55) | 1.32 (-2.90) | 1 | 11.28 (1.12–113.27) | 10.79 (1.17–99.50) | 3.06 (-0.15) |
| **Fatigue** | 6 | 0.11 (0.03–0.42) | 0.12 (0.04–0.42) | -0.62 (-2.08) | 0 | 5.94 (0.31–114.54) | 5.53 (0.36–84.05) | 2.41 (-1.83) | 2 | 6.66 (1.34–32.99) | 6.24 (1.39–28.00) | 2.38 (0.13) | 2 | 8.84 (1.76–44.47) | 8.10 (1.83–35.83) | 2.74 (0.47) |
| **Gastrointestinal disorder** | 4 | 0.16 (0.03–0.89) | 0.16 (0.03–0.87) | -0.47 (-2.29) | 0 | 9.66 (0.48–193.27) | 8.94 (0.56–141.46) | 3.06 (-1.23) | 0 | 1.87 (0.10–34.04) | 1.85 (0.11–32.11) | 0.84 (-3.33) | 2 | 17.79 (3.07–103.15) | 16.19 (3.14–83.54) | 3.48 (1.09) |
| **Headache** | 4 | 0.16 (0.03–0.89) | 0.16 (0.03–0.87) | -0.47 (-2.29) | 1 | 34.55 (3.26–366.40) | 27.84 (3.93–197.37) | 4.55 (1.25) | 1 | 5.15 (0.58–45.64) | 4.99 (0.60–41.28) | 2.11 (-0.98) | 0 | 2.41 (0.13–44.23) | 2.38 (0.14–40.96) | 1.19 (-2.99) |

| | | | | | | | | | | | | | | | | |
|---|---|---|---|---|---|---|---|---|---|---|---|---|---|---|---|---|
| **Hepatic function abnormal** | 2 | 0.08 (0.01–0.57) | 0.08 (0.01–0.57) | -0.89 (-3.33) | 0 | 13.99 (0.67–292.43) | 12.91 (0.78–214.20) | 3.55 (-0.80) | 2 | 26.88 (3.64–198.67) | 24.96 (3.65–170.59) | 3.70 (1.20) | 0 | 3.50 (0.18–67.01) | 3.44 (0.19–61.93) | 1.67 (-2.56) |
| **Hyperactivity*** | 3 | 0.24 (0.02–2.37) | 0.25 (0.03–2.32) | -0.30 (-2.46) | 0 | 13.99 (0.67–292.43) | 12.91 (0.78–214.20) | 3.55 (-0.80) | 1 | 8.60 (0.87–85.54) | 8.32 (0.89–77.41) | 2.70 (-0.50) | 0 | 3.50 (0.18–67.01) | 3.44 (0.19–61.93) | 1.67 (-2.56) |
| **Hyperglycaemia** | 0 | 0.01 (0.00–0.22) | 0.01 (0.00–0.23) | -2.89 (-7.12) | 0 | 18.01 (0.83–391.83) | 16.60 (0.96–287.53) | 3.87 (-0.52) | 2 | 53.84 (4.72–613.69) | 49.93 (4.67–533.80) | 4.11 (1.48) | 1 | 16.95 (1.48–194.73) | 16.19 (1.53–171.63) | 3.48 (0.16) |
| **Hyperphagia** | 5 | 0.40 (0.05–3.53) | 0.41 (0.05–3.44) | -0.15 (-1.86) | 0 | 9.66 (0.48–193.27) | 8.94 (0.56–141.46) | 3.06 (-1.23) | 1 | 5.15 (0.58–45.64) | 4.99 (0.60–41.28) | 2.11 (-0.98) | 0 | 2.41 (0.13–44.23) | 2.38 (0.14–40.96) | 1.19 (-2.99) |
| **Hyperprolactinaemia** | 2 | 0.16 (0.01–1.81) | 0.16 (0.02–1.77) | -0.47 (-3.05) | 0 | 18.01 (0.83–391.83) | 16.60 (0.96–287.53) | 3.87 (-0.52) | 1 | 12.92 (1.14–147.12) | 12.48 (1.17–133.45) | 3.11 (-0.19) | 0 | 4.50 (0.23–89.89) | 4.42 (0.24–83.04) | 1.99 (-2.28) |
| **Hypotension** | 0 | 0.03 (0.00–0.67) | 0.03 (0.00–0.67) | -1.89 (-6.35) | 0 | 42.15 (1.54–1152.96) | 38.72 (1.75–857.96) | 4.87 (0.26) | 0 | 8.16 (0.33–205.00) | 8.04 (0.33–192.90) | 2.65 (-1.85) | 1 | 99.59 (3.94–2519.20) | 92.86 (3.89–2216.12) | 4.58 (1.48) |
| **Insomnia** | 1 | 0.25 (0.01–6.16) | 0.25 (0.01–6.05) | -0.30 (-3.35) | 0 | 42.15 (1.54–1152.96) | 38.72 (1.75–857.96) | 4.87 (0.26) | 0 | 8.16 (0.33–205.00) | 8.04 (0.33–192.90) | 2.65 (-1.85) | 0 | 10.53 (0.42–266.14) | 10.32 (0.43–246.24) | 2.99 (-1.51) |
| **Lethargy** | 1 | 0.08 (0.00–1.30) | 0.08 (0.01–1.29) | -0.89 (-4.35) | 0 | 25.25 (1.08–589.47) | 23.23 (1.24–434.30) | 4.28 (-0.18) | 0 | 4.89 (0.23–104.34) | 4.82 (0.24–98.09) | 2.06 (-2.28) | 1 | 33.95 (2.05–562.39) | 32.38 (2.10–500.30) | 4.06 (0.55) |
| **Mania*** | 0 | 0.03 (0.00–0.67) | 0.03 (0.00–0.67) | -1.89 (-6.35) | 0 | 42.15 (1.54–1152.96) | 38.72 (1.75–857.96) | 4.87 (0.26) | 1 | 76.36 (3.04–1918.99) | 72.32 (3.01–1736.08) | 4.23 (1.14) | 0 | 10.53 (0.42–266.14) | 10.32 (0.43–246.24) | 2.99 (-1.51) |
| **Menstruation irregular** | 0 | 0.03 (0.00–0.67) | 0.03 (0.00–0.67) | -1.89 (-6.35) | 0 | 42.15 (1.54–1152.96) | 38.72 (1.75–857.96) | 4.87 (0.26) | 0 | 8.16 (0.33–205.00) | 8.04 (0.33–192.90) | 2.65 (-1.85) | 1 | 99.59 (3.94–2519.20) | 92.86 (3.89–2216.12) | 4.58 (1.48) |
| **Myalgia** | 1 | 0.08 (0.00–1.30) | 0.08 (0.01–1.29) | -0.89 (-4.35) | 0 | 25.25 (1.08–589.47) | 23.23 (1.24–434.30) | 4.28 (-0.18) | 1 | 25.88 (1.57–425.41) | 24.96 (1.60–388.55) | 3.70 (0.20) | 0 | 6.31 (0.29–135.51) | 6.19 (0.31–125.17) | 2.41 (-1.94) |

| | | | | | | | | | | | | | | | | |
|---|---|---|---|---|---|---|---|---|---|---|---|---|---|---|---|---|
| **Nausea** | 1 | 0.08 (0.00–1.30) | 0.08 (0.01–1.29) | -0.89 (-4.35) | 1 | 173.75 (9.18–3289.72) | 139.20 (10.05–1928.46) | 6.13 (2.45) | 0 | 4.89 (0.23–104.34) | 4.82 (0.24–98.09) | 2.06 (-2.28) | 0 | 6.31 (0.29–135.51) | 6.19 (0.31–125.17) | 2.41 (-1.94) |
| **Neutropenia** | 0 | 0.01 (0.00–0.16) | 0.01 (0.00–0.17) | -3.21 (-7.40) | 0 | 13.99 (0.67–292.43) | 12.91 (0.78–214.20) | 3.55 (-0.80) | 4 | 258.32 (13.51–4938.41) | 216.96 (11.97–3932.27) | 4.49 (2.59) | 0 | 3.50 (0.18–67.01) | 3.44 (0.19–61.93) | 1.67 (-2.56) |
| **Osteoporosis*** | 1 | 0.25 (0.01–6.16) | 0.25 (0.01–6.05) | -0.30 (-3.35) | 0 | 42.15 (1.54–1152.96) | 38.72 (1.75–857.96) | 4.87 (0.26) | 0 | 8.16 (0.33–205.00) | 8.04 (0.33–192.90) | 2.65 (-1.85) | 0 | 10.53 (0.42–266.14) | 10.32 (0.43–246.24) | 2.99 (-1.51) |
| **Pain** | 0 | 0.02 (0.00–0.34) | 0.02 (0.00–0.34) | -2.47 (-6.79) | 0 | 25.25 (1.08–589.47) | 23.23 (1.24–434.30) | 4.28 (-0.18) | 1 | 25.88 (1.57–425.41) | 24.96 (1.60–388.55) | 3.70 (0.20) | 1 | 33.95 (2.05–562.39) | 32.38 (2.10–500.30) | 4.06 (0.55) |
| **Panic attack*** | 0 | 0.03 (0.00–0.67) | 0.03 (0.00–0.67) | -1.89 (-6.35) | 0 | 42.15 (1.54–1152.96) | 38.72 (1.75–857.96) | 4.87 (0.26) | 1 | 76.36 (3.04–1918.99) | 72.32 (3.01–1736.08) | 4.23 (1.14) | 0 | 10.53 (0.42–266.14) | 10.32 (0.43–246.24) | 2.99 (-1.51) |
| **Rash** | 2 | 0.16 (0.01–1.81) | 0.16 (0.02–1.77) | -0.47 (-3.05) | 0 | 18.01 (0.83–391.83) | 16.60 (0.96–287.53) | 3.87 (-0.52) | 1 | 12.92 (1.14–147.12) | 12.48 (1.17–133.45) | 3.11 (-0.19) | 0 | 4.50 (0.23–89.89) | 4.42 (0.24–83.04) | 1.99 (-2.28) |
| **Sedation** | 1 | 0.08 (0.00–1.30) | 0.08 (0.01–1.29) | -0.89 (-4.35) | 0 | 25.25 (1.08–589.47) | 23.23 (1.24–434.30) | 4.28 (-0.18) | 1 | 25.88 (1.57–425.41) | 24.96 (1.60–388.55) | 3.70 (0.20) | 0 | 6.31 (0.29–135.51) | 6.19 (0.31–125.17) | 2.41 (-1.94) |
| **Sleep attacks*** | 1 | 0.25 (0.01–6.16) | 0.25 (0.01–6.05) | -0.30 (-3.35) | 0 | 42.15 (1.54–1152.96) | 38.72 (1.75–857.96) | 4.87 (0.26) | 0 | 8.16 (0.33–205.00) | 8.04 (0.33–192.90) | 2.65 (-1.85) | 0 | 10.53 (0.42–266.14) | 10.32 (0.43–246.24) | 2.99 (-1.51) |
| **Somnambulation*** | 1 | 0.25 (0.01–6.16) | 0.25 (0.01–6.05) | -0.30 (-3.35) | 0 | 42.15 (1.54–1152.96) | 38.72 (1.75–857.96) | 4.87 (0.26) | 0 | 8.16 (0.33–205.00) | 8.04 (0.33–192.90) | 2.65 (-1.85) | 0 | 10.53 (0.42–266.14) | 10.32 (0.43–246.24) | 2.99 (-1.51) |
| **Somnolence** | 576 | 76.80 (29.61–199.20) | 9.42 (4.09–21.71) | 0.10 (0.03) | 0 | 0.02 (0.00–0.33) | 0.10 (0.01–1.42) | -3.32 (-7.47) | 0 | 0.00 (0.00–0.05) | 0.02 (0.00–0.32) | -5.54 (-9.57) | 5 | 0.06 (0.02–0.16) | 0.28 (0.13–0.60) | -1.80 (-3.20) |
| **Urinary retention** | 1 | 0.25 (0.01–6.16) | 0.25 (0.01–6.05) | -0.30 (-3.35) | 0 | 42.15 (1.54–1152.96) | 38.72 (1.75–857.96) | 4.87 (0.26) | 0 | 8.16 (0.33–205.00) | 8.04 (0.33–192.90) | 2.65 (-1.85) | 0 | 10.53 (0.42–266.14) | 10.32 (0.43–246.24) | 2.99 (-1.51) |

| | | | | | | | | | | | | | | | | |
|---|---|---|---|---|---|---|---|---|---|---|---|---|---|---|---|---|
| **Vomiting** | 0 | 0.03 (0.00–0.67) | 0.03 (0.00–0.67) | -1.89 (-6.35) | 1 | 464.33 (16.58–13001.11) | 348.50 (15.73–7721.63) | 6.45 (3.20) | 0 | 8.16 (0.33–205.00) | 8.04 (0.33–192.90) | 2.65 (-1.85) | 0 | 10.53 (0.42–266.14) | 10.32 (0.43–246.24) | 2.99 (-1.51) |
| **Weight loss** | 1 | 0.25 (0.01–6.16) | 0.25 (0.01–6.05) | -0.30 (-3.35) | 0 | 42.15 (1.54–1152.96) | 38.72 (1.75–857.96) | 4.87 (0.26) | 0 | 8.16 (0.33–205.00) | 8.04 (0.33–192.90) | 2.65 (-1.85) | 0 | 10.53 (0.42–266.14) | 10.32 (0.43–246.24) | 2.99 (-1.51) |

### Supplementary Table 2: Gender-related differences in SOC-level of ADR reporting associated with atypical antipsychotics (AAPs) in paediatric patients with autism spectrum disorder (ASD)(OR, 95% CI).

| Drug | SOC | Male ADR no. | Female ADR no. | OR (95% CI) | P value |
|---|---|---|---|---|---|
| Olanzapine | Blood and lymphatic system disorders | 4 | 0 | 0.10 (0.00–2.11) | 0.11 |
| Quetiapine | Cardiac disorders | 3 | 1 | 0.18 (0.02–2.15) | 0.27 |
| Risperidone | Endocrine disorders | 2 | 1 | 0.77 (0.07–8.59) | 1.00 |
| Olanzapine | Endocrine disorders | 0 | 1 | 4.04 (0.15–108.57) | 0.21 |
| Risperidone | Gastrointestinal disorders | 6 | 7 | 1.83 (0.61–5.52) | 0.27 |
| Quetiapine | Gastrointestinal disorders | 1 | 1 | 0.73 (0.04–13.45) | 1.00 |
| Aripiprazole | Gastrointestinal disorders | 1 | 2 | 2.00 (0.05–78.25) | 1.00 |
| Risperidone | General disorders and administration site conditions | 3 | 3 | 1.56 (0.31–7.78) | 0.68 |
| Quetiapine | General disorders and administration site conditions | 1 | 2 | 1.60 (0.12–20.99) | 1.00 |
| Olanzapine | General disorders and administration site conditions | 1 | 2 | 2.80 (0.22–35.29) | 0.57 |
| Risperidone | Hepatobiliary disorders | 1 | 1 | 1.55 (0.10–24.95) | 1.00 |
| Olanzapine | Hepatobiliary disorders | 1 | 1 | 1.27 (0.07–22.72) | 1.00 |
| Risperidone | Metabolism and nutrition disorders | 6 | 8 | 2.10 (0.72–6.13) | 0.16 |
| Quetiapine | Metabolism and nutrition disorders | 0 | 1 | 2.48 (0.09–68.14) | 0.49 |
| Olanzapine | Metabolism and nutrition disorders | 2 | 3 | 2.17 (0.30–15.71) | 0.63 |
| Risperidone | Musculoskeletal and connective tissue disorders | 3 | 1 | 0.52 (0.05–4.98) | 1.00 |
| Olanzapine | Musculoskeletal and connective tissue disorders | 1 | 0 | 0.39 (0.01–10.37) | 0.49 |
| Risperidone | Nervous system disorders | 367 | 230 | 0.71 (0.40–1.25) | 0.23 |
| Quetiapine | Nervous system disorders | 3 | 6 | 2.00 (0.33–11.97) | 0.66 |
| Olanzapine | Nervous system disorders | 2 | 1 | 0.59 (0.05–7.43) | 1.00 |
| Aripiprazole | Nervous system disorders | 1 | 1 | 0.50 (0.01–19.56) | 1.00 |
| Risperidone | Psychiatric disorders | 3 | 2 | 1.03 (0.17–6.23) | 1.00 |
| Olanzapine | Psychiatric disorders | 3 | 2 | 0.80 (0.11–5.77) | 1.00 |
| Risperidone | Renal and urinary disorders | 1 | 0 | 0.52 (0.02–12.70) | 0.52 |
| Quetiapine | Reproductive system and breast disorders | 0 | 1 | 2.48 (0.09–68.14) | 0.49 |
| Olanzapine | Respiratory, thoracic and mediastinal disorders | 1 | 0 | 0.39 (0.01–10.37) | 0.49 |
| Risperidone | Skin and subcutaneous tissue disorders | 2 | 1 | 0.77 (0.07–8.59) | 1.00 |
| Olanzapine | Skin and subcutaneous tissue disorders | 0 | 2 | 7.38 (0.32–169.82) | 0.17 |
| Quetiapine | Vascular disorders | 1 | 0 | 0.23 (0.01–6.25) | 0.19 |

**Supplementary Table 3: Firth penalized logistic regression analysis of SOC-level ADR reporting associated with atypical antipsychotics (AAPs) in paediatric patients with autism spectrum disorders (ASD): adjusted odds ratios (aOR, 95%CI).** statistically significant findings

| AAP | SOC | Variable | Model used | Adjusted OR (95%CI) | P_value |
|---|---|---|---|---|---|
| **Olanzapine** | **Blood and lymphatic system disorders** | GenderFemale | Gender only_Firth | 0.1 (0–1.13) | 0.07 |
| **Quetiapine** | **Cardiac disorders** | GenderFemale | Age+Gender_Firth | 0.19 (0.01–1.77) | 0.15 |
| **Quetiapine** | **Cardiac disorders** | AgeGroup10-19 | Age+Gender_Firth | 0.12 (0.01–1.1) | 0.06 |
| **Olanzapine** | **Endocrine disorders** | GenderFemale | Gender only_Firth | 4.04 (0.2–612.67) | 0.37 |
| **Risperidone** | **Endocrine disorders** | AgeGroup10-19 | Age+Gender_Firth | 4.45 (0.4–34.13) | 0.19 |
| **Risperidone** | **Endocrine disorders** | GenderFemale | Age+Gender_Firth | 1 (0.09–7.67) | 1.00 |
| **Quetiapine** | **Gastrointestinal disorders** | AgeGroup10-19 | Age+Gender_Firth | 0.5 (0.04–6.78) | 0.57 |
| **Quetiapine** | **Gastrointestinal disorders** | GenderFemale | Age+Gender_Firth | 0.75 (0.06–10.11) | 0.81 |
| **Risperidone** | **Gastrointestinal disorders** | AgeGroup10-19 | Age+Gender_Firth | 5.46 (1.69–16.31) | 0.01 |
| **Risperidone** | **Gastrointestinal disorders** | GenderFemale | Age+Gender_Firth | 2.03 (0.69–6.17) | 0.20 |
| **Olanzapine** | **General disorders and administration site conditions** | GenderFemale | Gender only_Firth | 2.3 (0.27–27.98) | 0.45 |
| **Quetiapine** | **General disorders and administration site conditions** | AgeGroup10-19 | Age+Gender_Firth | 0.88 (0.1–10.73) | 0.91 |
| **Quetiapine** | **General disorders and administration site conditions** | GenderFemale | Age+Gender_Firth | 1.33 (0.15–16) | 0.79 |
| **Risperidone** | **General disorders and administration site conditions** | AgeGroup10-19 | Age+Gender_Firth | 14.97 (3.23–87.84) | <0.001 |
| **Risperidone** | **General disorders and administration site conditions** | GenderFemale | Age+Gender_Firth | 1.94 (0.4–9.53) | 0.40 |
| **Olanzapine** | **Hepatobiliary disorders** | GenderFemale | Gender only_Firth | 1.26 (0.09–17.15) | 0.85 |
| **Risperidone** | **Hepatobiliary disorders** | GenderFemale | Age+Gender_Firth | 1.79 (0.14–22.5) | 0.62 |
| **Risperidone** | **Hepatobiliary disorders** | AgeGroup10-19 | Age+Gender_Firth | 7.9 (0.63–99.3) | 0.10 |
| **Olanzapine** | **Metabolism and nutrition disorders** | GenderFemale | Gender only_Firth | 1.99 (0.32–14.1) | 0.46 |
| **Quetiapine** | **Metabolism and nutrition disorders** | AgeGroup10-19 | Age+Gender_Firth | 1.7 (0.08–260.21) | 0.75 |
| **Quetiapine** | **Metabolism and nutrition disorders** | GenderFemale | Age+Gender_Firth | 2.43 (0.12–367.31) | 0.57 |
| **Risperidone** | **Metabolism and nutrition disorders** | GenderFemale | Age+Gender_Firth | 2.08 (0.74–6.14) | 0.16 |
| **Risperidone** | **Metabolism and nutrition disorders** | AgeGroup10-19 | Age+Gender_Firth | 1.61 (0.31–5.55) | 0.52 |
| **Olanzapine** | **Musculoskeletal and connective tissue disorders** | GenderFemale | Gender only_Firth | 0.39 (0–7.94) | 0.55 |
| **Risperidone** | **Musculoskeletal and connective tissue disorders** | AgeGroup10-19 | Age+Gender_Firth | 17.42 (2.81–181.95) | <0.001 |
| **Risperidone** | **Musculoskeletal and connective tissue disorders** | GenderFemale | Age+Gender_Firth | 0.82 (0.08–5.19) | 0.84 |
| **Olanzapine** | **Nervous system disorders** | GenderFemale | Gender only_Firth | 0.7 (0.06–6.1) | 0.75 |
| **Quetiapine** | **Nervous system disorders** | GenderFemale | Age+Gender_Firth | 1.86 (0.35–11.1) | 0.47 |
| **Quetiapine** | **Nervous system disorders** | AgeGroup10-19 | Age+Gender_Firth | 2.18 (0.38–15.45) | 0.39 |
| **Risperidone** | **Nervous system disorders** | AgeGroup10-19 | Age+Gender_Firth | 0.15 (0.08–0.29) | <0.001 |
| **Risperidone** | **Nervous system disorders** | GenderFemale | Age+Gender_Firth | 0.61 (0.34–1.11) | 0.10 |
| **Olanzapine** | **Psychiatric disorders** | GenderFemale | Gender only_Firth | 0.85 (0.12–5.28) | 0.86 |
| **Risperidone** | **Psychiatric disorders** | AgeGroup10-19 | Age+Gender_Firth | 2.51 (0.25–13.89) | 0.37 |
| **Risperidone** | **Psychiatric disorders** | GenderFemale | Age+Gender_Firth | 1.14 (0.19–5.94) | 0.87 |
| **Risperidone** | **Renal and urinary disorders** | AgeGroup10-19 | Age+Gender_Firth | 20.79 (1.09–3055.9) | 0.04 |
| **Risperidone** | **Renal and urinary disorders** | GenderFemale | Age+Gender_Firth | 0.67 (0–12.98) | 0.80 |
| **Quetiapine** | **Reproductive system and breast disorders** | AgeGroup10-19 | Age+Gender_Firth | 1.7 (0.08–260.21) | 0.75 |
| **Quetiapine** | **Reproductive system and breast disorders** | GenderFemale | Age+Gender_Firth | 2.43 (0.12–367.31) | 0.57 |
| **Olanzapine** | **Respiratory, thoracic and mediastinal disorders** | GenderFemale | Gender only_Firth | 0.39 (0–7.94) | 0.55 |

| | | | | | |
|---|---|---|---|---|---|
| **Olanzapine** | **Skin and subcutaneous tissue disorders** | GenderFemale | Gender only_Firth | 7.38 (0.53–1059.33) | 0.15 |
| **Risperidone** | **Skin and subcutaneous tissue disorders** | AgeGroup10-19 | Age+Gender_Firth | 12.64 (1.65–140.18) | 0.02 |
| **Risperidone** | **Skin and subcutaneous tissue disorders** | GenderFemale | Age+Gender_Firth | 1.13 (0.1–8.74) | 0.91 |
| **Quetiapine** | **Vascular disorders** | GenderFemale | Age+Gender_Firth | 0.24 (0–4.83) | 0.35 |
| **Quetiapine** | **Vascular disorders** | AgeGroup10-19 | Age+Gender_Firth | 1.75 (0.08–276.71) | 0.73 |

**Supplementary Table 4: Gender-related differences in PT-level ADR reporting associated with atypical antipsychotics (AAPs)in paediatric patients with autism spectrum disorder (ASD) (OR, 95% CI).**

| AAP | PT | Male ADR no. | Female ADR no. | OR (95% CI) | P value |
|---|---|---|---|---|---|
| Risperidone | Abdominal distension | 0 | 1 | 4.67 (0.19-115.06) | 0.15 |
| Risperidone | Abdominal pain | 2 | 1 | 0.77 (0.07-8.59) | 1.00 |
| Risperidone | Abnormal weight gain | 3 | 5 | 2.62 (0.62-11.05) | 0.27 |
| Olanzapine | Abnormal weight gain | 0 | 2 | 7.38 (0.32-169.82) | 0.17 |
| Olanzapine | Alopecia | 0 | 1 | 4.04 (0.15-108.57) | 0.21 |
| Risperidone | Back pain | 2 | 0 | 0.31 (0.01-6.45) | 0.52 |
| Olanzapine | Confusional state | 2 | 0 | 0.22 (0.01-4.95) | 0.49 |
| Risperidone | Constipation | 1 | 1 | 1.55 (0.10-24.95) | 1.00 |
| Aripiprazole | Constipation | 1 | 0 | 0.14 (0.00-5.95) | 0.43 |
| Olanzapine | Cough | 1 | 0. | 0.39 (0.01-10.37) | 0.49 |
| Risperidone | Diarrhea | 1 | 0 | 0.52 (0.02-12.70) | 0.52 |
| Risperidone | Dizziness | 9 | 2 | 0.34 (0.07-1.58) | 0.22 |
| Quetiapine | Dizziness | 1 | 1 | 0.73 (0.04-13.45) | 1.00 |
| Olanzapine | Dizziness | 1 | 0 | 0.39 (0.01-10.37) | 0.49 |
| Aripiprazole | Dizziness | 1 | 0 | 0.14 (0.00-5.95) | 0.43 |
| Risperidone | Dry mouth | 0 | 1 | 4.67 (0.19-115.06) | 0.15 |
| Quetiapine | Electrocardiogram QT prolonged | 3 | 1 | 0.18 (0.02-2.15) | 0.27 |
| Risperidone | Endocrine disorder | 1 | 0 | 0.52 (0.02-12.70) | 0.52 |
| Risperidone | Erythema | 1 | 0 | 0.52 (0.02-12.70) | 0.52 |
| Risperidone | Extrapyramidal disorder | 2 | 1 | 0.77 (0.07-8.59) | 1.00 |
| Quetiapine | Extrapyramidal disorder | 0 | 1 | 2.48 (0.09-68.14) | 0.49 |
| Risperidone | Fatigue | 3 | 3 | 1.56 (0.31-7.78) | 0.68 |
| Quetiapine | Fatigue | 0 | 2 | 4.52 (0.19-106.70) | 0.48 |
| Olanzapine | Fatigue | 1 | 1 | 1.27 (0.07-22.72) | 1.00 |
| Risperidone | Gastrointestinal disorder | 1 | 3 | 4.70 (0.49-45.41) | 0.31 |
| Quetiapine | Gastrointestinal disorder | 1 | 1 | 0.73 (0.04-13.45) | 1.00 |
| Risperidone | Headache | 2 | 2 | 1.56 (0.22-11.11) | 0.65 |
| Olanzapine | Headache | 1 | 0 | 0.39 (0.01-10.37) | 0.49 |
| Aripiprazole | Headache | 0 | 1 | 3.00 (0.08-115.35) | 0.47 |
| Risperidone | Hepatic function abnormal | 1 | 1 | 1.55 (0.10-24.95) | 1.00 |
| Olanzapine | Hepatic function abnormal | 1 | 1 | 1.27 (0.07-22.72) | 1.00 |
| Risperidone | Hyperactivity | 2 | 1 | 0.77 (0.07-8.59) | 1.00 |
| Olanzapine | Hyperactivity | 1 | 0 | 0.39 (0.01-10.37) | 0.49 |
| Quetiapine | Hyperglycaemia | 0 | 1 | 2.48 (0.09-68.14) | 0.49 |
| Olanzapine | Hyperglycaemia | 2 | 0 | 0.22 (0.01-4.95) | 0.49 |
| Risperidone | Hyperphagia | 3 | 2 | 1.03 (0.17-6.23) | 1.00 |
| Olanzapine | Hyperphagia | 0 | 1 | 4.04 (0.15-108.57) | 0.21 |
| Risperidone | Hyperprolactinaemia | 1 | 1 | 1.55 (0.10-24.95) | 1.00 |
| Olanzapine | Hyperprolactinaemia | 0 | 1 | 4.04 (0.15-108.57) | 0.21 |
| Quetiapine | Hypotension | 1 | 0 | 0.23 (0.01-6.25) | 0.19 |
| Risperidone | Insomnia | 1 | 0 | 0.52 (0.02-12.70) | 0.52 |
| Risperidone | Lethargy | 1 | 0 | 0.52 (0.02-12.70) | 0.52 |

| | | | | | |
|---|---|---|---|---|---|
| **Quetiapine** | **Lethargy** | 0 | 1 | 2.48 (0.09-68.14) | 0.49 |
| **Olanzapine** | **Mania** | 0 | 1 | 4.04 (0.15-108.57) | 0.21 |
| **Quetiapine** | **Menstruation irregular** | 0 | 1 | 2.48 (0.09-68.14) | 0.49 |
| **Risperidone** | **Myalgia** | 0 | 1 | 4.67 (0.19-115.06) | 0.15 |
| **Olanzapine** | **Myalgia** | 1 | 0 | 0.39 (0.01-10.37) | 0.49 |
| **Risperidone** | **Nausea** | 1 | 0 | 0.52 (0.02-12.70) | 0.52 |
| **Aripiprazole** | **Nausea** | 0 | 1 | 3.00 (0.08-115.35) | 0.47 |
| **Olanzapine** | **Neutropenia** | 4 | 0 | 0.10 (0.00-2.11) | 0.11 |
| **Risperidone** | **Osteoporosis** | 1 | 0 | 0.52 (0.02-12.70) | 0.52 |
| **Quetiapine** | **Pain** | 1 | 0 | 0.23 (0.01-6.25) | 0.19 |
| **Olanzapine** | **Pain** | 0 | 1 | 4.04 (0.15-108.57) | 0.21 |
| **Olanzapine** | **Panic attack** | 0 | 1 | 4.04 (0.15-108.57) | 0.21 |
| **Risperidone** | **Rash** | 1 | 1 | 1.55 (0.10-24.95) | 1.00 |
| **Olanzapine** | **Rash** | 0 | 1 | 4.04 (0.15-108.57) | 0.21 |
| **Risperidone** | **Sedation** | 1 | 0 | 0.52 (0.02-12.70) | 0.52 |
| **Olanzapine** | **Sedation** | 0 | 1 | 4.04 (0.15-108.57) | 0.21 |
| **Risperidone** | **Sleep attacks** | 1 | 0 | 0.52 (0.02-12.70) | 0.52 |
| **Risperidone** | **Somnambulation** | 0 | 1 | 4.67 (0.19-115.06) | 0.15 |
| **Risperidone** | **Somnolence** | 351 | 225 | 0.95 (0.58-1.57) | 0.84 |
| **Quetiapine** | **Somnolence** | 2 | 3 | 1.17 (0.15-9.01) | 1.00 |
| **Risperidone** | **Urinary retention** | 1 | 0 | 0.52 (0.02-12.70) | 0.52 |
| **Aripiprazole** | **Vomiting** | 0 | 1 | 3.00 (0.08-115.35) | 0.47 |
| **Risperidone** | **Weight loss** | 0 | 1 | 4.67 (0.19-115.06) | 0.15 |

**Supplementary Table 5: Firth penalized logistic regression analysis of PT-level ADR reporting associated with atypical antipsychotics (AAPs) in paediatric with autism spectrum disorders (ASD): (aOR, 95%CI).**

| AAP | PT | Variable | Model used | Adjusted OR (95%CI) | P value |
|---|---|---|---|---|---|
| **Risperidone** | **Abdominal distension** | GenderFemale | Age+Gender_Firth | 4.7 (0.25–689.27) | 0.30 |
| **Risperidone** | **Abdominal distension** | AgeGroup10-19 | Age+Gender_Firth | 2.95 (0.02–57.12) | 0.55 |
| **Risperidone** | **Abdominal pain** | AgeGroup10-19 | Age+Gender_Firth | 4.45 (0.4–34.13) | 0.19 |
| **Risperidone** | **Abdominal pain** | GenderFemale | Age+Gender_Firth | 1 (0.09–7.67) | 1.00 |
| **Olanzapine** | **Abnormal weight gain** | GenderFemale | Gender only_Firth | 7.38 (0.53–1059.33) | 0.15 |
| **Risperidone** | **Abnormal weight gain** | GenderFemale | Age+Gender_Firth | 2.61 (0.68–11.38) | 0.16 |
| **Risperidone** | **Abnormal weight gain** | AgeGroup10-19 | Age+Gender_Firth | 3.2 (0.58–12.95) | 0.16 |
| **Olanzapine** | **Alopecia** | GenderFemale | Gender only_Firth | 4.04 (0.2–612.67) | 0.37 |
| **Risperidone** | **Back pain** | GenderFemale | Age+Gender_Firth | 0.35 (0–4.37) | 0.45 |
| **Risperidone** | **Back pain** | AgeGroup10-19 | Age+Gender_Firth | 6.74 (0.54–84.02) | 0.12 |
| **Olanzapine** | **Confusional state** | GenderFemale | Gender only_Firth | 0.22 (0–3) | 0.28 |
| **Risperidone** | **Constipation** | AgeGroup10-19 | Age+Gender_Firth | 7.9 (0.63–99.3) | 0.10 |
| **Risperidone** | **Constipation** | GenderFemale | Age+Gender_Firth | 1.79 (0.14–22.5) | 0.62 |
| **Olanzapine** | **Cough** | GenderFemale | Gender only_Firth | 0.39 (0–7.94) | 0.55 |
| **Risperidone** | **Diarrhea** | GenderFemale | Age+Gender_Firth | 0.51 (0–9.71) | 0.67 |
| **Risperidone** | **Diarrhea** | AgeGroup10-19 | Age+Gender_Firth | 2.25 (0.02–42.83) | 0.65 |
| **Olanzapine** | **Dizziness** | GenderFemale | Gender only_Firth | 0.39 (0–7.94) | 0.55 |
| **Quetiapine** | **Dizziness** | AgeGroup10-19 | Age+Gender_Firth | 2.9 (0.21–411.89) | 0.46 |
| **Quetiapine** | **Dizziness** | GenderFemale | Age+Gender_Firth | 0.74 (0.05–10.53) | 0.81 |
| **Risperidone** | **Dizziness** | AgeGroup10-19 | Age+Gender_Firth | 30.26 (8.35–160.84) | <0.001 |
| **Risperidone** | **Dizziness** | GenderFemale | Age+Gender_Firth | 0.49 (0.09–1.85) | 0.31 |
| **Risperidone** | **Dry mouth** | GenderFemale | Age+Gender_Firth | 6.39 (0.33–949.39) | 0.22 |
| **Risperidone** | **Dry mouth** | AgeGroup10-19 | Age+Gender_Firth | 28.02 (1.45–4148.5) | 0.03 |
| **Quetiapine** | **Electrocardiogram QT prolonged** | GenderFemale | Age+Gender_Firth | 0.19 (0.01–1.77) | 0.15 |
| **Quetiapine** | **Electrocardiogram QT prolonged** | AgeGroup10-19 | Age+Gender_Firth | 0.12 (0.01–1.1) | 0.06 |
| **Risperidone** | **Endocrine disorder** | AgeGroup10-19 | Age+Gender_Firth | 20.79 (1.09–3055.9) | 0.04 |
| **Risperidone** | **Endocrine disorder** | GenderFemale | Age+Gender_Firth | 0.67 (0–12.98) | 0.80 |
| **Risperidone** | **Erythema** | AgeGroup10-19 | Age+Gender_Firth | 20.79 (1.09–3055.9) | 0.04 |
| **Risperidone** | **Erythema** | GenderFemale | Age+Gender_Firth | 0.67 (0–12.98) | 0.80 |
| **Quetiapine** | **Extrapyramidal disorder** | GenderFemale | Age+Gender_Firth | 2.66 (0.12–431.81) | 0.55 |
| **Quetiapine** | **Extrapyramidal disorder** | AgeGroup10-19 | Age+Gender_Firth | 0.16 (0–3.29) | 0.23 |
| **Risperidone** | **Extrapyramidal disorder** | AgeGroup10-19 | Age+Gender_Firth | 12.64 (1.65–140.18) | 0.02 |
| **Risperidone** | **Extrapyramidal disorder** | GenderFemale | Age+Gender_Firth | 1.13 (0.1–8.74) | 0.91 |
| **Olanzapine** | **Fatigue** | GenderFemale | Gender only_Firth | 1.26 (0.09–17.15) | 0.85 |
| **Quetiapine** | **Fatigue** | GenderFemale | Age+Gender_Firth | 4.66 (0.32–687.71) | 0.29 |
| **Quetiapine** | **Fatigue** | AgeGroup10-19 | Age+Gender_Firth | 3.24 (0.21–485.77) | 0.44 |
| **Risperidone** | **Fatigue** | AgeGroup10-19 | Age+Gender_Firth | 14.97 (3.23–87.84) | <0.001 |
| **Risperidone** | **Fatigue** | GenderFemale | Age+Gender_Firth | 1.94 (0.4–9.53) | 0.40 |
| **Quetiapine** | **Gastrointestinal disorder** | AgeGroup10-19 | Age+Gender_Firth | 0.5 (0.04–6.78) | 0.58 |
| **Quetiapine** | **Gastrointestinal disorder** | GenderFemale | Age+Gender_Firth | 0.75 (0.06–10.11) | 0.82 |
| **Risperidone** | **Gastrointestinal disorder** | GenderFemale | Age+Gender_Firth | 3.87 (0.63–40.33) | 0.15 |
| **Risperidone** | **Gastrointestinal disorder** | AgeGroup10-19 | Age+Gender_Firth | 3.69 (0.35–23.23) | 0.23 |
| **Olanzapine** | **Headache** | GenderFemale | Gender only_Firth | 0.39 (0–7.94) | 0.55 |

| | | | | | |
|---|---|---|---|---|---|
| **Risperidone** | **Headache** | AgeGroup10-19 | Age+Gender_Firth | 76.05 (7.9–10154.32) | <0.001 |
| **Risperidone** | **Headache** | GenderFemale | Age+Gender_Firth | 2.22 (0.33–15.25) | 0.39 |
| **Olanzapine** | **Hepatic function abnormal** | GenderFemale | Gender only_Firth | 1.26 (0.09–17.15) | 0.85 |
| **Risperidone** | **Hepatic function abnormal** | AgeGroup10-19 | Age+Gender_Firth | 7.9 (0.63–99.3) | 0.10 |
| **Risperidone** | **Hepatic function abnormal** | GenderFemale | Age+Gender_Firth | 1.79 (0.14–22.5) | 0.62 |
| **Olanzapine** | **Hyperactivity** | GenderFemale | Gender only_Firth | 0.39 (0–7.94) | 0.55 |
| **Risperidone** | **Hyperactivity** | GenderFemale | Age+Gender_Firth | 0.9 (0.08–6.8) | 0.92 |
| **Risperidone** | **Hyperactivity** | AgeGroup10-19 | Age+Gender_Firth | 1.04 (0.01–10.91) | 0.98 |
| **Olanzapine** | **Hyperglycaemia** | GenderFemale | Gender only_Firth | 0.22 (0–3) | 0.28 |
| **Quetiapine** | **Hyperglycaemia** | GenderFemale | Age+Gender_Firth | 2.43 (0.12–367.31) | 0.57 |
| **Quetiapine** | **Hyperglycaemia** | AgeGroup10-19 | Age+Gender_Firth | 1.7 (0.08–260.21) | 0.75 |
| **Olanzapine** | **Hyperphagia** | GenderFemale | Gender only_Firth | 4.04 (0.2–612.67) | 0.37 |
| **Risperidone** | **Hyperphagia** | AgeGroup10-19 | Age+Gender_Firth | 0.67 (0.01–6.05) | 0.78 |
| **Risperidone** | **Hyperphagia** | GenderFemale | Age+Gender_Firth | 1.07 (0.18–5.52) | 0.94 |
| **Olanzapine** | **Hyperprolactinaemia** | GenderFemale | Gender only_Firth | 4.04 (0.2–612.67) | 0.37 |
| **Risperidone** | **Hyperprolactinaemia** | GenderFemale | Age+Gender_Firth | 1.51 (0.12–18.75) | 0.72 |
| **Risperidone** | **Hyperprolactinaemia** | AgeGroup10-19 | Age+Gender_Firth | 1.54 (0.01–19.32) | 0.79 |
| **Quetiapine** | **Hypotension** | GenderFemale | Age+Gender_Firth | 0.24 (0–4.83) | 0.35 |
| **Quetiapine** | **Hypotension** | AgeGroup10-19 | Age+Gender_Firth | 1.75 (0.08–276.71) | 0.73 |
| **Risperidone** | **Insomnia** | AgeGroup10-19 | Age+Gender_Firth | 20.79 (1.09–3055.9) | 0.04 |
| **Risperidone** | **Insomnia** | GenderFemale | Age+Gender_Firth | 0.67 (0–12.98) | 0.80 |
| **Quetiapine** | **Lethargy** | GenderFemale | Age+Gender_Firth | 2.66 (0.12–431.81) | 0.55 |
| **Quetiapine** | **Lethargy** | AgeGroup10-19 | Age+Gender_Firth | 0.16 (0–3.29) | 0.23 |
| **Risperidone** | **Lethargy** | AgeGroup10-19 | Age+Gender_Firth | 20.79 (1.09–3055.9) | 0.04 |
| **Risperidone** | **Lethargy** | GenderFemale | Age+Gender_Firth | 0.67 (0–12.98) | 0.80 |
| **Olanzapine** | **Mania** | GenderFemale | Gender only_Firth | 4.04 (0.2–612.67) | 0.37 |
| **Quetiapine** | **Menstruation irregular** | GenderFemale | Age+Gender_Firth | 2.43 (0.12–367.31) | 0.57 |
| **Quetiapine** | **Menstruation irregular** | AgeGroup10-19 | Age+Gender_Firth | 1.7 (0.08–260.21) | 0.75 |
| **Olanzapine** | **Myalgia** | GenderFemale | Gender only_Firth | 0.39 (0–7.94) | 0.55 |
| **Risperidone** | **Myalgia** | GenderFemale | Age+Gender_Firth | 6.39 (0.33–949.39) | 0.22 |
| **Risperidone** | **Myalgia** | AgeGroup10-19 | Age+Gender_Firth | 28.02 (1.45–4148.5) | 0.03 |
| **Risperidone** | **Nausea** | AgeGroup10-19 | Age+Gender_Firth | 20.79 (1.09–3055.9) | 0.04 |
| **Risperidone** | **Nausea** | GenderFemale | Age+Gender_Firth | 0.67 (0–12.98) | 0.80 |
| **Olanzapine** | **Neutropenia** | GenderFemale | Gender only_Firth | 0.1 (0–1.13) | 0.07 |
| **Risperidone** | **Osteoporosis** | AgeGroup10-19 | Age+Gender_Firth | 20.79 (1.09–3055.9) | 0.04 |
| **Risperidone** | **Osteoporosis** | GenderFemale | Age+Gender_Firth | 0.67 (0–12.98) | 0.80 |
| **Olanzapine** | **Pain** | GenderFemale | Gender only_Firth | 4.04 (0.2–612.67) | 0.37 |
| **Quetiapine** | **Pain** | GenderFemale | Age+Gender_Firth | 0.21 (0–4.89) | 0.34 |
| **Quetiapine** | **Pain** | AgeGroup10-19 | Age+Gender_Firth | 0.15 (0–3.3) | 0.23 |
| **Olanzapine** | **Panic attack** | GenderFemale | Gender only_Firth | 4.04 (0.2–612.67) | 0.37 |
| **Olanzapine** | **Rash** | GenderFemale | Gender only_Firth | 4.04 (0.2–612.67) | 0.37 |
| **Risperidone** | **Rash** | AgeGroup10-19 | Age+Gender_Firth | 7.9 (0.63–99.3) | 0.10 |
| **Risperidone** | **Rash** | GenderFemale | Age+Gender_Firth | 1.79 (0.14–22.5) | 0.62 |
| **Olanzapine** | **Sedation** | GenderFemale | Gender only_Firth | 4.04 (0.2–612.67) | 0.37 |
| **Risperidone** | **Sedation** | GenderFemale | Age+Gender_Firth | 0.51 (0–9.71) | 0.67 |
| **Risperidone** | **Sedation** | AgeGroup10-19 | Age+Gender_Firth | 2.25 (0.02–42.83) | 0.65 |
| **Risperidone** | **Sleep attacks** | AgeGroup10-19 | Age+Gender_Firth | 20.79 (1.09–3055.9) | 0.04 |

| | | | | | |
|---|---|---|---|---|---|
| **Risperidone** | **Sleep attacks** | GenderFemale | Age+Gender_Firth | 0.67 (0–12.98) | 0.80 |
| **Risperidone** | **Somnambulation** | GenderFemale | Age+Gender_Firth | 4.7 (0.25–689.27) | 0.30 |
| **Risperidone** | **Somnambulation** | AgeGroup10-19 | Age+Gender_Firth | 2.95 (0.02–57.12) | 0.55 |
| **Quetiapine** | **Somnolence** | AgeGroup10-19 | Age+Gender_Firth | 8.09 (0.75–1108.66) | 0.09 |
| **Quetiapine** | **Somnolence** | GenderFemale | Age+Gender_Firth | 1.13 (0.15–9.48) | 0.90 |
| **Risperidone** | **Somnolence** | AgeGroup10-19 | Age+Gender_Firth | 0.07 (0.04–0.12) | <0.001 |
| **Risperidone** | **Somnolence** | GenderFemale | Age+Gender_Firth | 0.77 (0.44–1.34) | 0.35 |
| **Risperidone** | **Urinary retention** | AgeGroup10-19 | Age+Gender_Firth | 20.79 (1.09–3055.9) | 0.04 |
| **Risperidone** | **Urinary retention** | GenderFemale | Age+Gender_Firth | 0.67 (0–12.98) | 0.80 |
| **Risperidone** | **Weight loss** | GenderFemale | Age+Gender_Firth | 4.7 (0.25–689.27) | 0.30 |
| **Risperidone** | **Weight loss** | AgeGroup10-19 | Age+Gender_Firth | 2.95 (0.02–57.12) | 0.55 |

**Supplementary Table 6: Age-group differences in SOC level ADR reporting associated with atypical antipsychotics (AAPs) in paediatric patients with autism spectrum disorder (ASD) (OR, 95% CI).**

statistically significant findings

| Drug | SOC | Children (0-9) ADR no. | Adolescents (10-19) ADR no. | OR (95% CI) | P value |
|---|---|---|---|---|---|
| Olanzapine | Blood and lymphatic system disorders | 0 | 4 | 0.19 (0.00-10.97) | 1.00 |
| Quetiapine | Cardiac disorders | 3 | 1 | 0.10 (0.01-1.28) | 0.09 |
| Risperidone | Endocrine disorders | 2 | 1 | 3.80 (0.34-42.42) | 0.31 |
| Olanzapine | Endocrine disorders | 0 | 2 | 0.06 (0.00-3.97) | 1.00 |
| Risperidone | Gastrointestinal disorders | 8 | 5 | 4.96 (1.58-15.59) | 0.01 |
| Quetiapine | Gastrointestinal disorders | 1 | 1 | 0.46 (0.02-8.69) | 1.00 |
| Aripiprazole | Gastrointestinal disorders | 1 | 2 | 2.00 (0.05-78.25) | 1.00 |
| Risperidone | General disorders and administration site conditions | 2 | 4 | 15.83 (2.85-87.98) | <0.001 |
| Quetiapine | General disorders and administration site conditions | 1 | 2 | 1.00 (0.07-13.37) | 1.00 |
| Olanzapine | General disorders and administration site conditions | 0 | 4 | 0.14 (0.00-8.42) | 1.00 |
| Risperidone | Hepatobiliary disorders | 1 | 1 | 7.61 (0.47-123.00) | 0.22 |
| Olanzapine | Hepatobiliary disorders | 0 | 2 | 0.10 (0.00-6.09) | 1.00 |
| Quetiapine | Metabolism and nutrition disorders | 0 | 2 | 1.67 (0.06-46.23) | 0.54 |
| Risperidone | Metabolism and nutrition disorders | 12 | 2 | 1.26 (0.28-5.75) | 0.67 |
| Olanzapine | Metabolism and nutrition disorders | 0 | 6 | 0.24 (0.00-13.74) | 1.00 |
| Risperidone | Musculoskeletal and connective tissue disorders | 1 | 3 | 23.47 (2.41-228.57) | 0.01 |
| Olanzapine | Musculoskeletal and connective tissue disorders | 0 | 2 | 0.06 (0.00-3.97) | 1.00 |
| Risperidone | Nervous system disorders | 541 | 56 | 0.16 (0.09-0.30) | <0.001 |
| Quetiapine | Nervous system disorders | 2 | 7 | 2.50 (0.36-17.50) | 0.64 |
| Olanzapine | Nervous system disorders | 0 | 4 | 0.14 (0.00-8.42) | 1.00 |
| Aripiprazole | Nervous system disorders | 1 | 1 | 0.50 (0.01-19.56) | 1.00 |
| Risperidone | Psychiatric disorders | 4 | 1 | 1.89 (0.21-17.17) | 0.47 |
| Olanzapine | Psychiatric disorders | 0 | 6 | 0.24 (0.00-13.74) | 1.00 |
| Risperidone | Renal and urinary disorders | 0 | 2 | 22.75 (0.92-563.49) | 0.01 |
| Quetiapine | Reproductive system and breast disorders | 0 | 2 | 1.67 (0.06-46.23) | 0.54 |
| Olanzapine | Respiratory, thoracic and mediastinal disorders | 0 | 2 | 0.06 (0.00-3.97) | 1.00 |
| Risperidone | Skin and subcutaneous tissue disorders | 1 | 2 | 15.43 (1.38-172.29) | 0.04 |
| Olanzapine | Skin and subcutaneous tissue disorders | 0 | 2 | 0.10 (0.00-6.09) | 1.00 |
| Quetiapine | Vascular disorders | 0 | 2 | 1.67 (0.06-46.23) | 0.54 |

### Supplementary Table 7: Age-group differences in PT-level ADR reporting associated with atypical antipsychotics (AAPs) in paeadtric patients with autism spectrum disorders (ASD) (OR, 95% CI).

| Drug | PT | Adolescents (10-19) ADR no. | Children (0-9) ADR no. | OR (95% CI) | P value |
|---|---|---|---|---|---|
| Risperidone | Abdominal distension | 0 | 1 | 2.49 (0.10-61.68) | 1.00 |
| Risperidone | Abdominal pain | 1 | 2 | 3.80 (0.34-42.42) | 0.31 |
| Risperidone | Abnormal weight gain | 2 | 6 | 2.55 (0.51-12.86) | 0.24 |
| Olanzapine | Abnormal weight gain | 0 | 2 | 10.20 (0.16-633.97) | 1.00 |
| Olanzapine | Alopecia | 0 | 1 | 17.67 (0.25-1239.98) | 1.00 |
| Risperidone | Back pain | 1 | 1 | 7.61 (0.47-123.00) | 0.22 |
| Olanzapine | Confusional state | 0 | 2 | 10.20 (0.16-633.97) | 1.00 |
| Risperidone | Constipation | 1 | 1 | 7.61 (0.47-123.00) | 0.22 |
| Aripiprazole | Constipation | 0 | 1 | 0.14 (0.00-5.95) | 0.43 |
| Olanzapine | Cough | 0 | 1 | 17.67 (0.25-1239.98) | 1.00 |
| Risperidone | Diarrhea | 0 | 1 | 2.49 (0.10-61.68) | 1.00 |
| Risperidone | Dizziness | 9 | 2 | 38.28 (8.10-180.91) | 0.00 |
| Quetiapine | Dizziness | 2 | 0 | 3.00 (0.13-71.32) | 0.52 |
| Olanzapine | Dizziness | 0 | 1 | 17.67 (0.25-1239.98) | 1.00 |
| Aripiprazole | Dizziness | 0 | 1 | 0.14 (0.00-5.95) | 0.43 |
| Risperidone | Dry mouth | 1 | 0 | 22.75 (0.92-563.49) | 0.01 |
| Quetiapine | Electrocardiogram QT prolonged | 1 | 3 | 0.10 (0.01-1.28) | 0.09 |
| Risperidone | Endocrine disorder | 1 | 0 | 22.75 (0.92-563.49) | 0.01 |
| Risperidone | Erythema | 1 | 0 | 22.75 (0.92-563.49) | 0.01 |
| Risperidone | Extrapyramidal disorder | 2 | 1 | 15.43 (1.38-172.29) | 0.04 |
| Quetiapine | Extrapyramidal disorder | 0 | 1 | 0.15 (0.01-4.18) | 0.12 |
| Risperidone | Fatigue | 4 | 2 | 15.83 (2.85-87.98) | 0.00 |
| Quetiapine | Fatigue | 2 | 0 | 3.00 (0.13-71.32) | 0.52 |
| Olanzapine | Fatigue | 0 | 2 | 10.20 (0.16-633.97) | 1.00 |
| Risperidone | Gastrointestinal disorder | 1 | 3 | 2.53 (0.26-24.63) | 0.39 |
| Quetiapine | Gastrointestinal disorder | 1 | 1 | 0.46 (0.02-8.69) | 1.00 |
| Risperidone | Headache | 4 | 0 | 71.07 (3.79-1333.61) | 0.00 |
| Olanzapine | Headache | 0 | 1 | 17.67 (0.25-1239.98) | 1.00 |
| Aripiprazole | Headache | 1 | 0 | 3.00 (0.08-115.35) | 0.47 |
| Risperidone | Hepatic function abnormal | 1 | 1 | 7.61 (0.47-123.00) | 0.22 |
| Olanzapine | Hepatic function abnormal | 0 | 2 | 10.20 (0.16-633.97) | 1.00 |
| Risperidone | Hyperactivity | 0 | 3 | 1.06 (0.05-20.79) | 1.00 |
| Olanzapine | Hyperactivity | 0 | 1 | 17.67 (0.25-1239.98) | 1.00 |
| Quetiapine | Hyperglycaemia | 1 | 0 | 1.67 (0.06-46.23) | 0.54 |
| Olanzapine | Hyperglycaemia | 0 | 2 | 10.20 (0.16-633.97) | 1.00 |
| Risperidone | Hyperphagia | 0 | 5 | 0.67 (0.04-12.32) | 1.00 |
| Olanzapine | Hyperphagia | 0 | 1 | 17.67 (0.25-1239.98) | 1.00 |

| | | | | | |
|---|---|---|---|---|---|
| **Risperidone** | **Hyperprolactinaemia** | 0 | 2 | 1.49 (0.07-31.36) | 1.00 |
| **Olanzapine** | **Hyperprolactinaemia** | 0 | 1 | 17.67 (0.25-1239.98) | 1.00 |
| **Quetiapine** | **Hypotension** | 1 | 0 | 1.67 (0.06-46.23) | 0.54 |
| **Risperidone** | **Insomnia** | 1 | 0 | 22.75 (0.92-563.49) | 0.01 |
| **Risperidone** | **Lethargy** | 1 | 0 | 22.75 (0.92-563.49) | 0.01 |
| **Quetiapine** | **Lethargy** | 0 | 1 | 0.15 (0.01-4.18) | 0.12 |
| **Olanzapine** | **Mania** | 0 | 1 | 17.67 (0.25-1239.98) | 1.00 |
| **Quetiapine** | **Menstruation irregular** | 1 | 0 | 1.67 (0.06-46.23) | 0.54 |
| **Risperidone** | **Myalgia** | 1 | 0 | 22.75 (0.92-563.49) | 0.01 |
| **Olanzapine** | **Myalgia** | 0 | 1 | 17.67 (0.25-1239.98) | 1.00 |
| **Risperidone** | **Nausea** | 1 | 0 | 22.75 (0.92-563.49) | 0.01 |
| **Aripiprazole** | **Nausea** | 1 | 0 | 3.00 (0.08-115.35) | 0.47 |
| **Risperidone** | **Osteoporosis** | 1 | 0 | 22.75 (0.92-563.49) | 0.01 |
| **Quetiapine** | **Pain** | 0 | 1 | 0.15 (0.01-4.18) | 0.12 |
| **Olanzapine** | **Pain** | 0 | 1 | 17.67 (0.25-1239.98) | 1.00 |
| **Olanzapine** | **Panic attack** | 0 | 1 | 17.67 (0.25-1239.98) | 1.00 |
| **Risperidone** | **Rash** | 1 | 1 | 7.61 (0.47-123.00) | 0.22 |
| **Olanzapine** | **Rash** | 0 | 1 | 17.67 (0.25-1239.98) | 1.00 |
| **Risperidone** | **Sedation** | 0 | 1 | 2.49 (0.10-61.68) | 1.00 |
| **Olanzapine** | **Sedation** | 0 | 1 | 17.67 (0.25-1239.98) | 1.00 |
| **Risperidone** | **Sleep attacks** | 1 | 0 | 22.75 (0.92-563.49) | 0.01 |
| **Risperidone** | **Somnambulation** | 0 | 1 | 2.49 (0.10-61.68) | 1.00 |
| **Risperidone** | **Somnolence** | 39 | 537 | 0.07 (0.04-0.12) | 0.00 |
| **Quetiapine** | **Somnolence** | 5 | 0 | 8.68 (0.41-183.24) | 0.07 |
| **Risperidone** | **Urinary retention** | 1 | 0 | 22.75 (0.92-563.49) | 0.01 |
| **Aripiprazole** | **Vomiting** | 1 | 0 | 3.00 (0.08-115.35) | 0.47 |
| **Risperidone** | **Weight loss** | 0 | 1 | 2.49 (0.10-61.68) | 1.00 |
